# Provisional research criteria for the behavioral variant of Alzheimer’s disease A systematic review and meta-analysis

**DOI:** 10.1101/2021.09.08.21263253

**Authors:** R. Ossenkoppele, E.H. Singleton, C. Groot, Anke A. Dijkstra, Willem S. Eikelboom, William W. Seeley, Bruce Miller, R. Laforce, P. Scheltens, J.M. Papma, G.D. Rabinovici, Y.A.L. Pijnenburg

**Affiliations:** Alzheimer Center Amsterdam, Department of Neurology, Amsterdam Neuroscience, Vrije Universiteit Amsterdam, Amsterdam UMC, Amsterdam, The Netherlands; Lund University, Clinical Memory Research Unit, Lund, Sweden; Department of Pathology, Amsterdam Neuroscience, Amsterdam University Medical Centre, Location VUMC; Department of Neurology, Erasmus University Medical Center, Rotterdam, the Netherlands; Memory and Aging Center, Department of Neurology, University of California San Francisco, San Francisco, USA; Clinique Interdisciplinaire de Mémoire (CIME), CHU de Québec, Québec, Canada; Department of Radiology and Biomedical Imaging, University of California San Francisco, San Francisco, USA; Weill Institute for Neurosciences, University of California San Francisco, San Francisco, CA, USA

## Abstract

**Importance:** The behavioral variant of Alzheimer’s disease (bvAD) is characterized by early and predominant behavioral deficits caused by AD pathology. This AD phenotype is insufficiently understood and lacks standardized clinical criteria, limiting reliability and reproducibility of diagnosis and scientific reporting.

**Objective:** To perform a systematic review and meta-analysis of the bvAD literature, and use the outcomes to propose provisional research criteria for this syndrome.

**Data sources:** A systematic literature search in PubMed/Medline and Web-of-Science databases (from inception through April 7th, 2021, performed in duplicate) led to the assessment of 83 studies, including 13 suitable for meta-analysis.

**Study selection:** Studies reporting on behavioral, neuropsychological or neuroimaging features in bvAD, and, when available, providing comparisons with “typical” amnestic-predominant AD (tAD) or behavorial variant frontotemporal dementia (bvFTD).

**Data extraction and synthesis:** We performed random-effects meta-analyses on group-level study results of clinical data, and systematically reviewed the neuroimaging literature.

**Main outcome and measures:** Behavioral symptoms (neuropsychiatric symptoms and bvFTD core clinical criteria), cognitive function (global cognition, episodic memory and executive functioning) and neuroimaging features (structural MRI, [^18^F]FDG-PET, perfusion SPECT, amyloid-PET and tau-PET).

**Results:** Data were collected for 591 patients with bvAD. There was moderate-to-substantial heterogeneity and moderate risk of bias across studies. bvAD showed more severe behavioral symptoms compared to tAD (standardized mean difference [SMD, 95% confidence interval]: 1.16[0.74–1.59], p<0.001), and a trend towards less severe behavioral symptoms compared to bvFTD (SMD:-0.22[-0.47–0.04], p=0.10). Meta-analyses of cognitive data indicated worse executive performance in bvAD versus tAD (SMD:-1.03[-1.74–-0.32], p<0.01), but not compared to bvFTD (SMD:-0.61[-1.75–0.53], p=0.29). bvAD showed a trend towards worse memory performance compared to bvFTD (SMD:-1.31[-2.75–0.14], p=0.08), but did not differ from tAD (SMD:0.43[-0.46–1.33], p=0.34). The neuroimaging literature revealed two distinct bvAD neuroimaging-phenotypes: an “AD-like” posterior-predominant pattern and a “bvFTD-like” anterior-predominant pattern, with the former being more prevalent.

**Conclusions and relevance:** Our data indicate that bvAD is clinically most similar to bvFTD, while it shares most pathophysiological features with tAD. Based on these insights, we propose provisional research criteria for bvAD aimed at improving the consistency and reliability of future research and aiding the clinical assessment of this AD phenotype.

**KEY POINTS:** *Question:* How does the behavioral variant of Alzheimer’s disease (bvAD) relate to typical AD (tAD) and to behavioral variant frontotemporal dementia (bvFTD) in terms of clinical presentation and neuroimaging signatures?

*Findings:* In this systematic review and meta-analysis, we found that, at time of diagnosis, bvAD showed more severe neuropsychiatric symptoms and other behavioral deficits compared to tAD. Two distinct neuroimaging phenotypes were observed across reported bvAD cases: an “AD-like” posterior-predominant pattern and a “bvFTD-like” anterior-predominant pattern, with the posterior-predominant neuroimaging phenotype being the most prevalent across reported bvAD cases.

*Meaning:* bvAD is clinically most reminiscent of bvFTD, while it shares most pathophysiological features with tAD. The provisional research criteria are aimed at improving the consistency and reliability of future research, and potentially aid in the clinical assessment of bvAD.

## INTRODUCTION

Alzheimer’s disease (AD) is a heterogenous disease that can manifest with both amnestic and non-amnestic clinical presentations.^1^ Several atypical (i.e., non-memory predominant) variants of AD have been described, including posterior cortical atrophy (PCA), logopenic variant primary progressive aphasia (lvPPA), corticobasal syndrome due-to-AD and dysexecutive AD.^2^ The behavioral variant of Alzheimer’s disease (bvAD) represents another, rare, variant of AD that is characterized by early and predominant behavioral deficits and personality changes caused by AD pathology. The bvAD clinical syndrome overlaps substantially with that of the behavioral variant of frontotemporal dementia (bvFTD) and ∼10-40% of clinically diagnosed bvFTD cases have positive AD biomarkers and/or neuropathologically confirmed AD.^3-6^ This highlights a major diagnostic challenge, which is even more pertinent with the recent accelerated approval of aducanumab by the FDA to reduce cerebral amyloid-β in early symptomatic AD.^7^ Although bvAD is acknowledged as a clinical entity in recent diagnostic/research criteria for AD dementia^8,9^, currently no criteria exist that provide specific recommendations for the diagnosis of bvAD. This is in contrast with other AD variants^10-12^, and limits reliable and reproducible classification of bvAD as well as uniform scientific reporting.

The current literature on bvAD comprises relatively few studies with typically small sample sizes that have reported several inconsistent findings. To better understand the bvAD phenotype, we performed a systematic review and meta-analysis of the clinical, neuroimaging, and neuropathology bvAD literature, and applied the outcomes to develop provisional research criteria for bvAD. With this work, we aim to improve the consistency and reliability of future research, and potentially aid in the clinical assessment of bvAD.

## METHODS

### Search strategy, selection and outcomes

This study was conducted following pre-specified methods (PROSPERO registration number: CRD42021243497) and reported following the PRISMA guidelines (**Table-S1**). We performed a systematic literature search in PubMed/Medline and Web-of-Science databases. We searched studies including clinically diagnosed 1) AD cases with “frontal” or “behavioral” presentations, or 2) bvFTD cases with neuropathological evidence of AD (see full database queries in **Table-S2**). We included peer-reviewed articles, written in English and presenting original research with human data only. Screening was first conducted at the title/abstract level in Rayyan (https://rayyan.qcri.org/). Reference lists were additionally cross-checked for eligible studies. Two independent reviewers (RO/EHS) screened titles and abstracts. Ambiguous records were discussed with a third author (YALP) to reach consensus. Studies were eligible when 1) including cases or groups of patients presenting with early and predominant behavioral changes with a clinical diagnosis, biomarker support and/or neuropathological evidence of AD, and 2) behavioral/neuropsychiatric, neuropsychological, neuroimaging and/or neuropathological data were presented. Studies were excluded when 1) describing patients with isolated executive dysfunction in the absence of behavioral symptoms, 2) there was biomarker and/or neuropathological evidence for a non-AD pathology as primary etiology. Studies were only eligible for the meta-analysis if a bvAD group was compared against typical AD (tAD) and/or bvFTD groups. We extracted demographic (age, sex), clinical (behavioral features per bvFTD criteria^13^ or neuropsychiatric symptoms per Neuropsychiatric Inventory [NPI^14^]), neuropsychological (Mini-Mental State Examination [MMSE], memory and executive function tests), neuroimaging (structural MRI, [^18^F]FDG-PET, perfusion SPECT, amyloid-PET, tau-PET) and neuropathological (amyloid-β and tau) characteristics from all studies. After eligibility assessment for inclusion, meta-analyses were constructed using pooled clinical (behavioral or neuropsychiatric questionnaires), neuropsychological (MMSE, memory and executive functioning tests) and neuropathological (amyloid-β and tau) data. The lack of uniform reporting of effect sizes among neuroimaging methods across studies did not allow a meta-analysis, hence these findings were analyzed using systematic review (**Table-1**).

**Table 1.** Neuroimaging studies in the behavioral variant of Alzheimer’s disease

| Study | Subjects | Age | Sex | MMSE | AD confirmation | Contrasts | Findings |
| --- | --- | --- | --- | --- | --- | --- | --- |
| **MRI** |  |  |  |  |  |  |  |
| Ossenkoppele et al. 2015<sup>23</sup> | 55 bvAD | 64.7<br>(8.8) | 72.7 | 22.5<br>(5.4) | CSF/PET/autopsy | 58 typical AD, 59 bvFTD, 61 CU | Predominant temporoparietal pattern, no differences between bvAD vs tAD |
| Perry et al. 2017*<sup>24</sup> | 15 bvAD | 62.8<br>(43-83) | 66.7 | n/a | Autopsy | 98 bvFTD | Moderate atrophy in frontoinsula and anterior cingulate regions |
| Phillips et al. 2018<sup>17</sup> | 22 b/dAD | 64.3<br>(8.2) | 50.0 | 19.6<br>(8.4) | CSF/autopsy | 22 typical AD, 115 CU | Staging scheme based on cross-sectional MRI data indicated early frontotemporal and insular atrophy and spread towards frontotemporal & parietal cortex |
| Phillips et al. 2019**<sup>25</sup> | 12 bvAD | 16.0<br>[13.5, 18.0] | 58.3 | 23.0<br>[17.0, 26.0] | CSF/autopsy | 17 typical AD | Anterior insula, frontotemporal, angular gyrus and middle temporal atrophy |
| Singleton et al. 2020*<sup>27</sup> | 29 bvAD | 64.4<br>(9.4) | 59.0 | 22.0<br>(5.9) | CSF/PET/autopsy | 28 typical AD, 28 bvFTD, 34 CU | Larger amygdalar grey matter volume in bvAD vs tAD |
| Therriault et al. 2020<sup>26</sup> | 15 b/dAD | 65.9<br>(8.8) | 40.0 | 19.6<br>(5.3) | Amyloid-and tau-PET positivity | 25 typical AD, 131 CU | Lateral and medial parietal/prefrontal atrophy in bvAD. No differences between bvAD vs tAD |
| **FDG-PET/Hypoperfusion SPECT** |  |  |  |  |  |  |  |
| Snowden et al. 2007<sup>36</sup> | 12 bvAD | 49 (8) | 75.0 | n/a | NA | 321 typical AD | Hypoperfusion extending into the frontal regions occurred more commonly in patients with "frontal" behavioral features |
| Woodward et al 2015<sup>30</sup> | 13 bvAD | 81.6<br>(4.1) | 61.5 | 23.9 | NA | 40 typical AD | Greater frontotemporal and parietal hypometabolism in bvAD vs tAD |
| Wang et al. 2019<sup>31</sup> | 13 b/dAD | 68.0<br>(3.4) | 30.8 | 17.0<br>(5.6) | Amyloid-PET positivity | 38 typical AD, 20 CU | Dorsolateral hypometabolism in the medial prefrontal and dorsolateral frontal cortex |
| Bergeron et al. 2020<sup>28</sup> | 8 b/dAD | 61.6 | n/a | 20.7 | CSF/PET | 40 atypical AD, 12 FTD | Most hypometabolic regions of interest in b/dAD were 1) middle temporal gyrus, 2) posterior cingulate, 3) angular gyrus |
| Singleton et al. 2020*<sup>27</sup> | 19 bvAD | 66.1<br>(7.4) | 58.0 | 21.8<br>(5.7) | CSF/PET/autopsy | 18 typical AD, 18 bvFTD, 31 CU | No hypometabolic differences in direct contrasts, but relatively greater frontoinsular involvement in bvAD vs tAD in contrasts again CU. |
| Sala et al. 2020<sup>29</sup> | 15 b/dAD | 62.5<br>(5.7) | 66.7 | 16.5<br>(5.2) | CSF | 22 typical AD | Temporoparietal, dorsolateral and orbitofrontal hypometabolism, highest (>90%) hypometabolism in the middle and superior frontal gyrus |
| Bergeron et al. 2021<sup>15</sup> | 6 b/dAD | 59.5<br>(7.9) | 75.0 | 22.3<br>(5.9) | CSF/PET | 8 bvFTD, 10 typical AD | 2/6 showed a predominantly frontal and temporoparietal pattern, 2/6 a mild frontal pattern, 2/6 temporoparietal predominant |
| Lehingue et al. 2021<sup>16</sup> | 20 bvAD | 71.5<br>[66-76] | 65.0 | 25 [21-26] | CSF | 22 typical AD, 36 bvFTD | No significant differences between bvAD vs tAD/bvFTD. |
| **Amyloid PET** |  |  |  |  |  |  |  |
| Wang et al. 2019<sup>31</sup> | 13 b/dAD | 68.0<br>(3.4) | 30.8 | 17.0<br>(5.6) | Amyloid-PET positivity | 38 typical AD, 20 CU | No differences among AD groups |
| Therriault et al. 2020<sup>26</sup> | 15 b/dAD | 65.9<br>(8.8) | 40.0 | 19.6<br>(5.3) | Amyloid- and tau-PET positivity | 25 typical AD, 131 CU | No differences among AD groups |

| Tau PET |  |  |  |  |  |  |  |
| --- | --- | --- | --- | --- | --- | --- | --- |
| Therriault et al. 2020<sup>26</sup> | 15 b/dAD | 65.9<br>(8.8) | 40.0 | 19.6<br>(5.3) | Amyloid- and tau-PET positivity | 25 typical AD, 131 CU | b/dAD vs tAD: elevated uptake in the anterior cingulate, medial prefrontal and frontal insula cortices in b/dAD vs tAD |
| Singleton et al. 2021<sup>32</sup> | 7 bvAD | 69.1<br>(8.4) | 85.7 | 21.7<br>(2.8) | CSF/PET/autopsy | 205 typical AD | 3/7 prominent lateral frontal and temporoparietal uptake, 1/7 medial prefrontal, 2/7 lateral temporal, 1/7 temporoparietal uptake. |

Values for age and MMSE represent the mean (sd), while for sex the percentage of male bvAD patients in the study are provided. *AD*=Alzheimer's disease, *bvAD*=behavioral variant of AD, *b/dAD*=behavioral/dysexecutive AD, *tAD*=typical AD, *CSF*=cerebrospinal fluid, *PET*=positron emission tomography, *MRI*=magnetic resonance imaging, *FDG*=fluorodeoxyglucose.
\*Cases partially overlap with Ossenkoppele et al. 2015
\*\*Cases partially overlap with Phillips et al. 2018

### Statistical analysis

Meta-analysis was used to examine whether bvAD differed from tAD and bvFTD in terms of behavioral/neuropsychiatric and neuropsychological features, and whether bvAD differed from tAD in the distribution of amyloid-β and tau pathology defined at autopsy. Missing data were requested from the authors of three studies (3/3 responded).^15-17^ We calculated the pooled standardized mean differences and 95% confidence intervals using Hedges’ g random-effects models in the “meta” package (R v4.0.2), with significance levels of p<0.05. We used random effects because we assumed that the true effect size would be study dependent, due to high heterogeneity in samples, methodology and outcomes among studies.

Statistical heterogeneity for the meta-analyses was assessed using the *I*^2^ statistic, with *I*^2^>75% indicating substantial heterogeneity. Heterogeneity across studies was substantial for analyses including behavioral/neuropsychiatric symptoms, memory and executive measures (range *I*^2^: 70-96%) and moderate for analyses including neuropathological data (range *I*^2^: 0-51%). Publication bias was assessed by visual inspection of funnel plots, which indicated substantial publication bias (**Figure-S1-S3**). Two authors (EHS/CG) independently assessed risk of bias using the ROBINS-I risk of bias tool for non-randomized studies. The overall risk of bias was serious for two studies and moderate for 11 studies in the meta-analysis (**Table-S3, Figure-S4**).

We additionally calculated the prevalence of each behavioral feature in the core clinical bvFTD criteria^13^ (i.e., presence of disinhibition, apathy, lack of empathy, compulsiveness and hyperorality) and the 12 items of the neuropsychiatric inventory (NPI, prevalence score ≥1) across studies, and compared bvAD, bvFTD and tAD groups using χ^2^ tests. For the NPI analysis only, we used 769 Aβ-positive tAD cases from the Amsterdam Dementia Cohort (mean age[SD]: 65.9[7.7], 52.4% females, mean[SD] MMSE: 20.3[5.1]).^18^

## RESULTS

### Participants

The systematic literature search yielded 1,257 records, of which 116 studies were assessed at full-text level for eligibility and 83 studies met inclusion criteria (see **Figure-S5** for flow chart). Thirteen studies were eligible for meta-analysis. **Table-S4** provides an overview of the participant characteristics for all 83 included studies. Across these studies, 591 bvAD cases were enrolled, with a mean(SD) age-at-diagnosis of 62.0(7.3), and (38.3%) of participants were female. The mean(SD) MMSE was 20.1(5.9) and 47.5% carried an *APOE*ε4 allele.

### Behavioral/neuropsychiatric symptoms

Meta-analysis indicated that patients with bvAD showed more severe behavioral/neuropsychiatric symptoms than tAD patients (standardized mean difference [SMD, 95% confidence interval]: 1.16[0.74–1.59], p<0.001), and a trend towards less severe behavioral/neuropsychiatric symptoms compared to bvFTD (SMD:-0.22[-0.47–0.04], p=0.10, **Figure-1A**). Results remained similar when separating bvFTD core criteria and neuropsychiatric features (**Figure-S6**).

**Figure 1.**
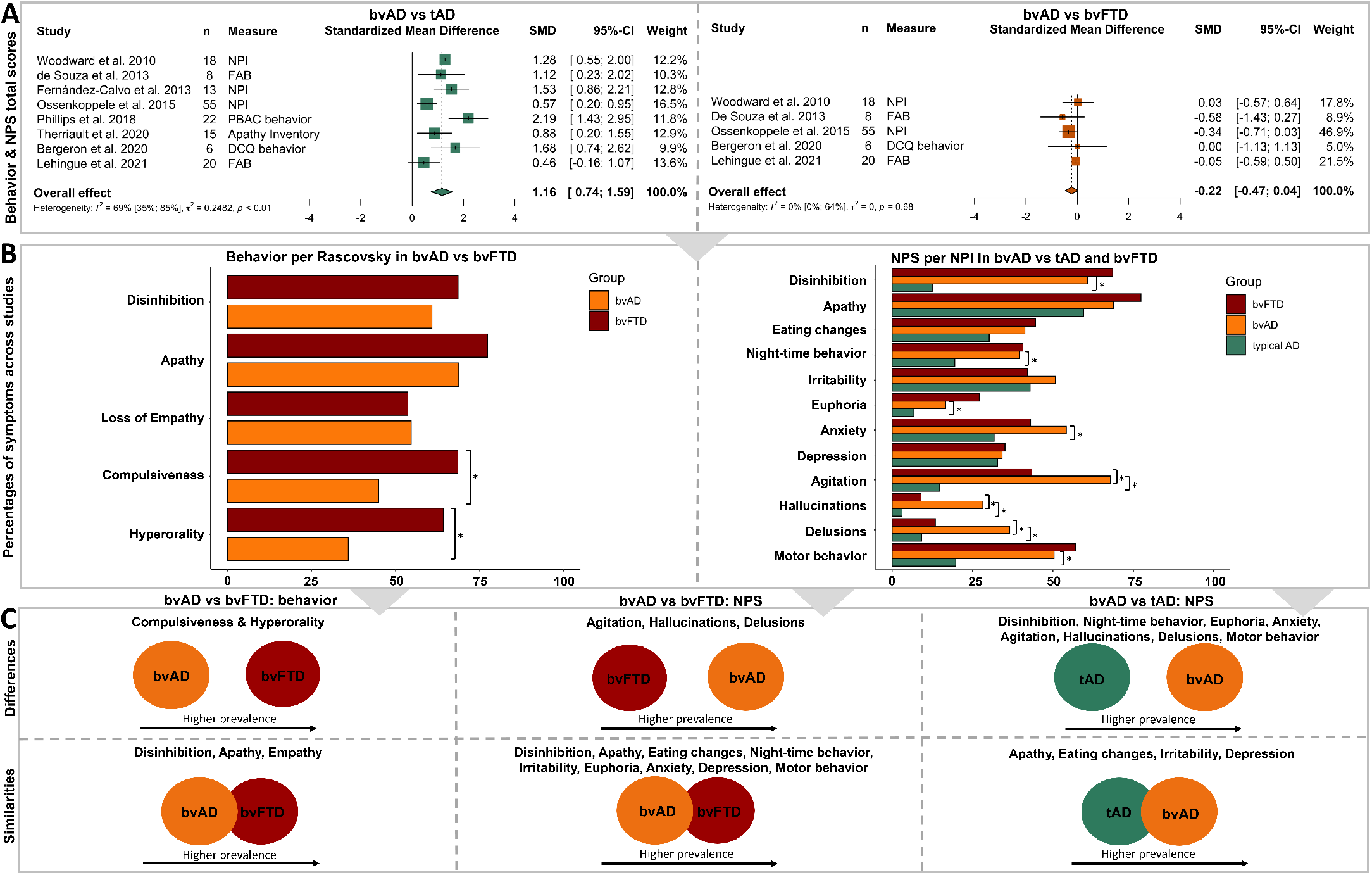
Meta-analyses of behavioral and neuropsychiatric features Panel A shows the results of meta-analyses on behavioral and neuropsychiatric total scores between bvAD and typical AD (left) and bvAD and bvFTD (right). “n” represents the number of bvAD cases in each study. Panel B shows the mean weighted percentages of participants per diagnostic group fulfilling specific bvFTD core clinical features proposed by Raskovsky et al. (left) or presence of specific neuropsychiatric symptoms measured using the Neuropsychiatric Inventory (NPI, right). Panel C summarizes the differences and similarities between diagnostic groups. *bvAD*=behavioral variant of AD, *tAD*=typical AD, *bvFTD*=behavioral variant frontotemporal dementia. *NPI*=Neuropsychiatric Inventory, *FAB*=Frontal Assessment Battery, *PBAC*=Philadelphia Brief Assessment of Cognition, *DCQ*=Dépistage Cognitif de Québec, *SMD*=standardized mean difference.

Next, we compared proportions of bvFTD features and NPI items as reported in previous studies (**Figure-1B, Table-S5**^19-25^). Compared to bvFTD, bvAD patients less frequently showed compulsive behaviors (45.0%*vs*68.5%, χ^2^=22.5, p<0.001) and hyperorality (35.9%*vs*64.1%, χ^2^=32.8, p<0.001), but no differences on disinhibition (60.8%*vs*68.6%, χ^2^=2.8, p=0.10), apathy (68.8%*vs*77.4%, χ^2^=3.7, p=0.05) and lack of empathy (54.6%*vs*53.6%, χ^2^=0.1, p=0.83). On the NPI, bvAD patients more frequently showed agitation (67.9*vs*43.4%, χ^2^=8.8), hallucinations (28.2*vs*9.0%, χ^2^=12.8) and delusions (36.6*vs*13.4%, χ^2^=13.4) compared to bvFTD (all p<0.001). Furthermore, bvAD more frequently showed night-time behaviors (39.6*vs*19.5%, χ^2^=12.9), euphoria (16.6*vs*6.8%, χ^2^=7.9), anxiety (54.2*vs*31.7%, χ^2^=10.8), agitation (67.9*vs*14.8%, χ^2^=90.3), hallucinations (28.2*vs*3.1%, χ^2^=71.2), delusions (36.6*vs*9.2%, χ^2^=37.2) and motor behaviors (50.4*vs*19.8%, χ^2^=26.2) compared to tAD patients (all p<0.01).

### Cognition

Meta-analyses of cognitive data indicated that at initial assessment bvAD patients showed no differences on MMSE compared to tAD (SMD:-0.18[-0.56–0.20], p=0.35) and bvFTD (SMD:-0.22[-0.78–0.35], p=0.46, **Figure-2**). bvAD showed worse executive performance compared to tAD (SMD:-1.03[-1.74–-0.32], p<0.01), but not compared to bvFTD (SMD:-0.61[-1.75–0.53], p=0.29). Finally, bvAD showed a trend towards worse memory performance compared to bvFTD (SMD:-1.31[-2.75–0.14], p=0.08), but did not differ from tAD (SMD:0.43[-0.46–1.33], p=0.34).

**Figure 2.**
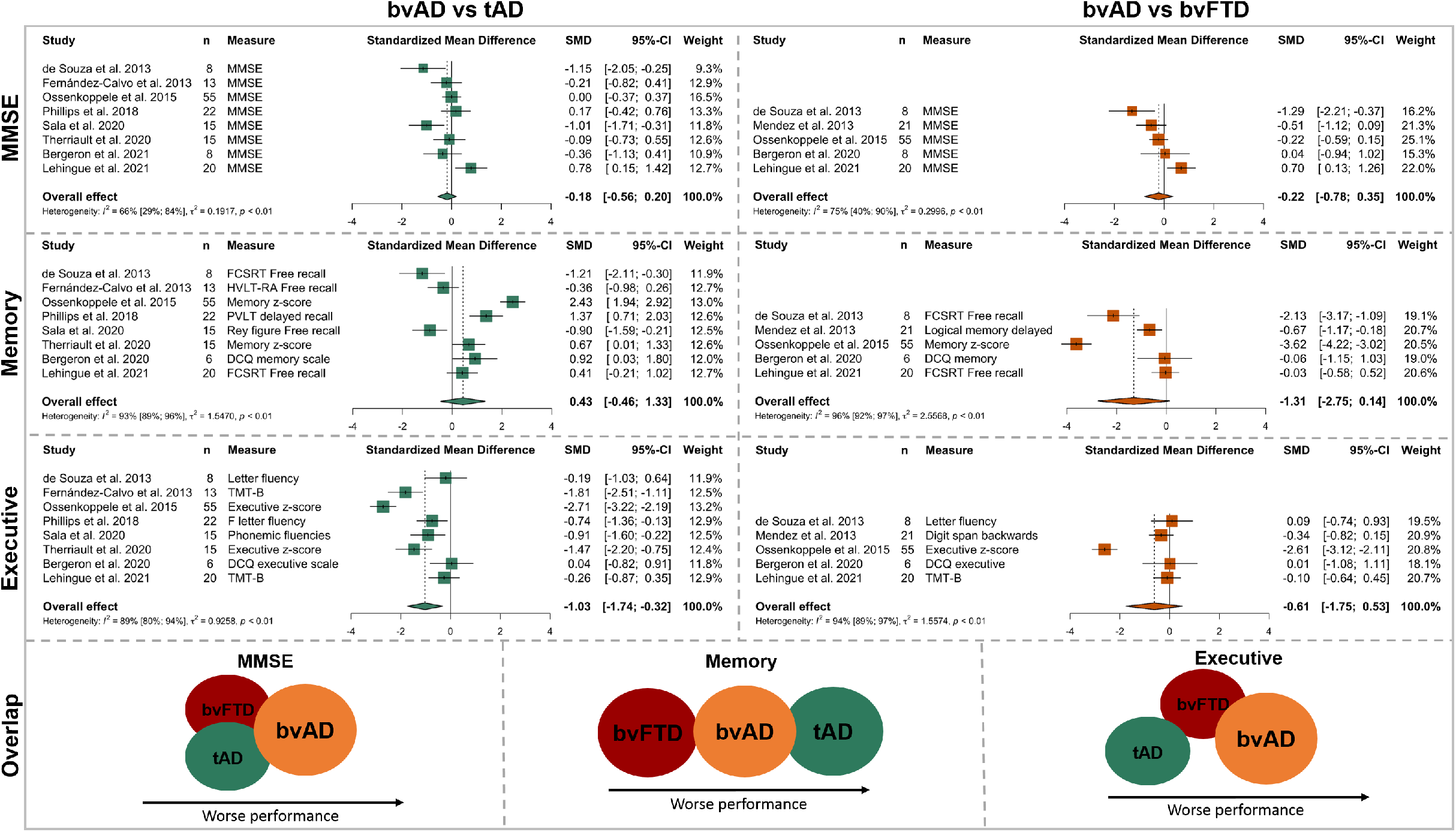
Meta-analyses of cognitive performance Results of meta-analyses on Mini-Mental State Examination, episodic memory and executive function for the contrast bvAD vs typical AD (left) and bvAD vs bvFTD (right). *bvAD*=behavioral variant of AD, *tAD*=typical AD, *bvFTD*=behavioral variant frontotemporal dementia. *SMD*=standardized mean difference.

### Neuroimaging

**Table-2** provides an overview of neuroimaging studies in bvAD. Structural MRI studies (number of studies [“k”]=6, number of participants [“n”]=92) showed temporoparietal^23^, frontotemporal and insular^17,24,25^, or frontoparietal^26^ predominant atrophy patterns across bvAD patients. bvAD did not differ from tAD in three studies^23,26,27^, and showed moderately more involvement of frontal regions in bvAD compared to tAD in three other studies^17,24,25^. Studies assessing glucose metabolism with [^18^F]FDG-PET or perfusion with SPECT (k=7, n=88) also showed heterogeneous results, ranging from a predominantly temporoparietal hypometabolic pattern^27,28^ to a mixed frontal and temporoparietal^15,16,29,30^ or predominantly frontal pattern^31^. Amyloid-PET studies (k=2, n=28) showed no differences in amyloid-β burden or distribution between bvAD and tAD patients.^26,31^ For tau-PET (k=2, n=22), one study showed a temporoparietal pattern with higher uptake in anterior regions in bvAD compared to tAD^26^, whereas another study showed heterogeneous patterns across bvAD patients^32^. Findings on functional connectivity (k=3, n=54) and white matter hyperintensities (k=1, n=29) in bvAD are presented in **Table-S6**.

**Table 2.** Provisional research criteria for the behavioral variant of Alzheimer’s disease

| I. Clinical bvAD |
| --- |
| (A) The clinical syndrome is characterized by:<br>I. Early, persistent, predominant, and progressive change or exacerbation of at least 2/5 core behavioral features of the diagnostic criteria for behavioral variant frontotemporal dementia (Rascovsky et al. [2011]<sup>13</sup>):<br>1. Behavioral disinhibition [one of the following symptoms (1.1–1.3) must be present]:<br>1.1 Socially inappropriate behavior<br>1.2 Loss of manners or decorum<br>1.3 Impulsive, rash or careless actions<br>2. Apathy or inertia [one of the following symptoms (2.1–2.2) must be present]:<br>2.1 Apathy<br>2.2 Inertia<br>3. Loss of empathy or sympathy [one of the following symptoms (3.1–3.2) must be present]:<br>3.1 Diminished response to other people’s needs and feelings<br>3.2 Diminished social interest, interrelatedness or personal warmth<br>4. Perseverative, stereotyped or compulsive/ritualistic behavior [one of the following symptoms (4.1–4.3) must be present]:<br>4.1 Simple repetitive movements<br>4.2 Complex, compulsive or ritualistic behaviors<br>4.3 Stereotypy of speech<br>5. Hyperorality and dietary changes [one of the following symptoms (5.1–5.3) must be present]:<br>5.1 Altered food preferences<br>5.2 Binge eating, increased consumption of alcohol or cigarettes<br>5.3 Oral exploration or consumption of inedible objects<br>II. <u>And</u>, documented impairment in executive functions and/or episodic memory with relatively preserved language and visuospatial abilities. |
| (B) Criteria for clinical bvAD are <u>not</u> met if the behavioral deficits are (better) accounted for by another concurrent (active) neurological (e.g., Lewy body dementia) or non-neurological medical (e.g., psychiatric) comorbidity, a known genetic mutation associated with familial bvFTD, or use of medication. |
| (C) Supportive features (not mandatory, A and B must be met):<br>1. Presence of hallucinations and/or delusions.<br>2. AD-specific (posterior-predominant temporoparietal pattern) and/or bvFTD-specific neuroimaging features (anterior-predominant frontotemporal pattern) on MRI, CT, perfusion SPECT and/or FDG-PET. |
| **II. Possible bvAD** |
| (A) Meets criteria for clinical bvAD (I), and |
| (B) There is *in vivo* biomarker evidence for the presence of amyloid-β pathology on amyloid PET and/or in CSF, and/or for the presence of tau pathology in CSF and/or plasma. |
| **III. Probable bvAD** |
| Meets criteria for clinical bvAD (I) or possible bvAD (II), with additional *in vivo* tau PET evidence for the presence of neocortical tau aggregates. |
| **IV. Definite bvAD** |
| (A) Meets criteria for clinical bvAD (I), possible bvAD (II) or probable bvAD (III), and |
| (B) Presence of AD is established by<br>1. Histopathological indication of AD as the primary etiology on biopsy or at autopsy, or<br>2. Presence of a known genetic mutation associated with familial AD. |

*AD*=Alzheimer's disease, *bvAD*=behavioral variant of Alzheimer's disease, *bvFTD*=behavioral variant of frontotemporal dementia, *MRI*=magnetic resonance imaging, *CT*=computed tomography, *SPECT*=single photon emission computed tomography, *FDG*=fluorodeoxyglucose, *PET*=positron emission tomography

We distilled two distinct bvAD neuroimaging-phenotypes from the literature, including a posterior-predominant (more “AD-like”) and an anterior-predominant (more “bvFTD-like”) phenotype (**Figure-3A**). We propose that these phenotypes occur on a continuum (**Figure-3B**), with the posterior-predominant phenotype being most prevalent (**Figure-3C**).

**Figure 3.**
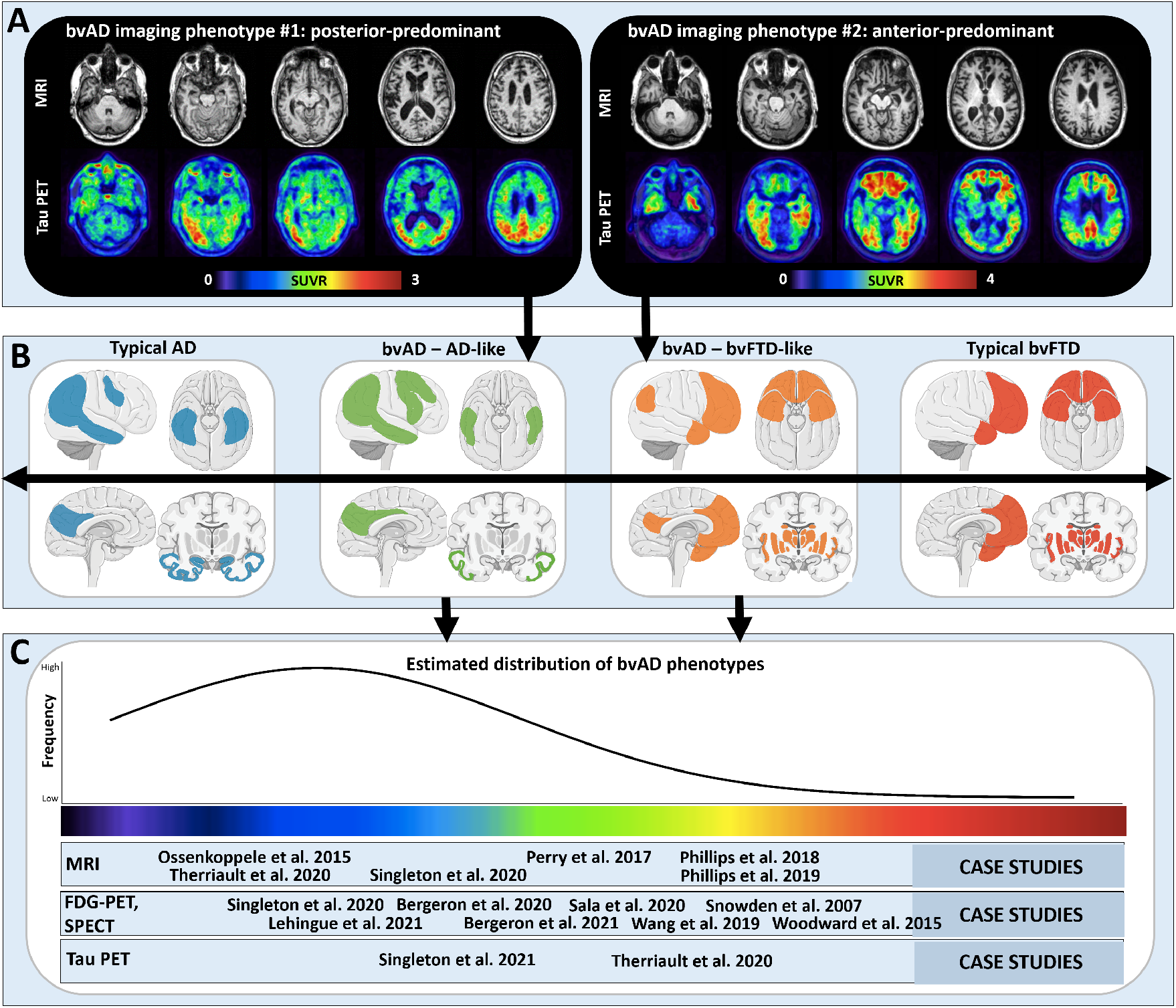
Neuroimaging features in bvAD The top panel (A) shows two cases which serve as examples of two distinct bvAD neuroimaging phenotypes: an “AD-like” posterior-predominant atrophy/tau load pattern and a “bvFTD-like” anterior-predominant atrophy/tau load pattern. In the middle panel (B) we propose that these neuroimaging phenotypes are part of a spectrum that ranges from a typical AD regional distribution to a classical bvFTD regional distribution (from left to right). The brain template images were obtained from https://smart.servier.com/. In the lower panel (C), we present a literature-informed estimated distribution of the regional distribution in bvAD, indicating that “typical AD” and “bvAD-AD-like” patterns are more common than “bvAD-bvFTD-like” and “typical bvFTD”.

### Neuropathology

In line with amyloid and tau PET findings, the meta-analyses on neuropathological data showed that bvAD and tAD did not differ in the neuropathological burden of amyloid-β (k=3, n=20) in the frontal cortex (SMD:0.23[-0.36–0.81], p=0.45), medial temporal lobe (SMD:-0.06[-0.65– 0.53], p=0.84) or occipital regions (SMD:-0.16[-1.05–0.73], p=0.73, **Figure-S7**). Furthermore, there was no difference in tau burden (k=4, n=28) in the frontal cortex (SMD:-0.05[-0.56–0.46], p=0.84), medial temporal lobe (SMD: 0.32[-0.19–0.83], p=0.22), or occipital lobe (SMD:-0.36[-0.95–0.23], p=0.24, **Figure-S7**).

## DISCUSSION

In this systematic review and meta-analysis, we found that bvAD is clinically most reminiscent of bvFTD, while it shares most pathophysiological features with tAD. Based on these insights, we provide provisional research criteria for bvAD aimed at improving the consistency and reliability of future research and aiding in future clinical assessments.

### Systematic review and meta-analyses

bvAD typically presents at a young age (62.0[7.3] years at time-of-diagnosis), is more prominent in males than in females (61.7%*vs*38.3%, in line with bvFTD but in contrast with tAD^33^) and has a lower frequency of *APOE*ε4 carriership compared to tAD (47.5%*vs*66.1%^34^). Clinically, bvAD shows a milder behavioral profile compared to bvFTD with less compulsivity and hyperorality, but greater prevalence of neuropsychiatric symptoms such as agitation, delusions and hallucinations. By definition, bvAD shows greater impairment on a range of behavioral and neuropsychiatric measures compared to tAD. The directionality of findings in the meta-analyses of cognitive data suggest that bvAD might show greater memory and executive function deficits compared to bvFTD, and relatively better memory function and worse executive functioning compared to tAD, but further research in larger cohorts is needed to confirm the significance of these findings. The neuroimaging methodology was too heterogenous across studies to conduct a formal meta-analysis, but a systematic review revealed two distinct phenotypes of brain atrophy, hypometabolism and tau pathology in bvAD, with many cases likely occurring on a continuum. The most prevalent bvAD neuroimaging phenotype is an “AD-like” posterior-predominant pattern comprising bilateral temporoparietal regions with limited involvement of the frontal cortex. This observation is congruent with our meta-analysis on neuropathological data showing that bvAD patients were indistinguishable from tAD patients in both amyloid-β and tau load and spatial distribution. The other bvAD phenotype is characterized by a “bvFTD-like” anterior-predominant neuroimaging pattern, including regions (e.g., anterior cingulate cortex, frontal insula, temporal poles) located in brain networks (e.g., the salience network) that are engaged during socio-emotional processing of information.^35^ Altogether, our systematic review and meta-analyses further refine the bvAD phenotype, but also highlight the need for larger studies with more uniform methodologies and inclusion/exclusion criteria.

### Provisional research criteria for bvAD

Our main objective was to propose provisional research criteria for bvAD guided by the results of the systematic review and meta-analyses. The criteria are based on consensus between all authors, consisting of neurologists, neuropsychologists, neuropathologists and neuroscientists. To facilitate widespread use but also take into account the complexity of this phenotype, we offer four levels of evidence (**Table-2**). The first level (“clinical bvAD”) can be established solely based on clinical information, while level II and III (“possible bvAD” and “probable bvAD”) add biomarker confirmation of amyloid-β and tau pathology. Level IV (“definite bvAD”) is assigned through histopathological or genetic (i.e., presence of pathogenic *APP, PSEN1* or *PSEN2* mutations) confirmation of AD in conjunction with a bvAD clinical syndrome.

Several issues warrant further explanation. First, both the literature and our clinical experience align with the notion that bvAD is a combined cognitive and behavioral clinical syndrome. We previously showed that cognitive impairment was among the first symptoms reported by patients and caregivers in ∼75% of bvAD cases.^23^ In addition, our meta-analysis suggests that episodic memory performance in bvAD is intermediate between tAD and bvFTD, while bvAD shows greater executive dysfunction compared to bvFTD (**Figure-1**). To enhance the discriminative accuracy between bvAD and bvFTD, objectively confirmed impairment in either memory or executive domains is therefore mandatory. In addition, 2/5 behavioral features of the diagnostic criteria for bvFTD^13^ (i.e., disinhibition, apathy, lack of empathy, compulsiveness and hyperorality) must be present. This 2:5 ratio was selected to sufficiently distinguish bvAD from tAD, but also acknowledge the generally milder behavioral profile in bvAD compared to bvFTD (where 3/6 bvFTD criteria must be present). Second, despite significant differences between bvAD and both bvFTD and tAD (**Figure-1**), we deemed it premature to include hallucinations and delusions in the core research criteria, as these observations were derived from only two studies.^21,22^ Instead, they were added as supportive features and future prospective studies are needed to assess whether they should be incorporated in the core criteria for bvAD. Third, most AD variants have a clear neurodegenerative signature on MRI and/or [^18^F]FDG-PET that corresponds to their clinical phenotype, such as left-hemispheric predominance in lvPPA or occipito-temporal/occipito-parietal damage in PCA.^10,11^ The neuroimaging literature in bvAD, however is highly inconsistent. Some studies (mainly case studies or case series) showed anterior neurodegenerative patterns that resemble bvFTD, but most group studies showed either a mix of anterior and posterior involvement or a posterior-predominant pattern.^17,23-28,36,37^ Contrary to PCA and lvPPA, we therefore did not incorporate MRI, CT, SPECT or [^18^F]FDG-PET read-outs into the core bvAD research criteria, but only added them as supportive features. Fourth, evidence of amyloid-β pathology provided by PET, CSF or plasma biomarkers can upgrade the diagnosis from “clinical bvAD” to “possible bvAD”. Positive amyloid-β biomarkers substantially increase the likelihood that AD is the primary etiology but, given their limited specificity, the possibility of amyloid-β as comorbid pathology cannot be ruled out, especially in older individuals and in *APOE*ε4 carriers.^38,39^ The addition of biomarker evidence for tau pathology further increases the certainty for a bvAD diagnosis (i.e., “probable bvAD”). Here we make the distinction between biofluid and neuroimaging markers of tau pathology. For CSF and plasma biomarkers of tau pathology the differential diagnostic value for distinguishing AD from bvFTD is less well established, and like amyloid-β markers they become abnormal relatively early in the disease course, which lowers their specificity.^40,41^ Hence, a full AD-like fluid biomarker profile with abnormalities in both amyloid-β and phosphorylated-tau supports a level II diagnosis of “possible bvAD”. Instead, the currently most widely used tau PET ligands (i.e., [^18^F]flortaucipir, [^18^F]MK6240 and [^18^F]RO948) have consistently shown to bind selectively and with high affinity to the tau aggregates formed in AD (i.e., combinations of 3R/4R tau in paired helical filaments), while neocortical tau PET uptake in sporadic bvFTD is negligible, resulting in excellent discriminative accuracy between AD and bvFTD.^42,43^ Furthermore, since tau PET-positivity in the neocortex almost exclusively occurs in amyloid-β positive individuals^42,44^ we consider a level I diagnosis (“clinical bvAD”) plus tau PET-positivity in an AD-like pattern^45^ supportive of a level III diagnosis of “probable bvAD”. Fifth, although the question whether bvAD and dysexecutive AD (deAD) reside on a single continuum or represent distinct clinical entities is yet unresolved, we deliberately developed criteria specific to bvAD. This was motivated by our previous study showing that only ∼25% of bvAD cases additionally met deAD criteria^23^ (hence bvAD occurs in isolation in the majority of cases), as well as a recent publication proposing specific deAD criteria that explicitly exclude behavioral features.^12^

### Limitations

There are several limitations. First, bvAD is a rare AD phenotype that, for the most part, has been described in single case studies and case series. The bvAD literature therefore consists of relatively few cohort studies that are generally characterized by modest sample sizes, which resulted in reduced statistical power to detect differences between bvAD versus bvFTD and tAD groups. This was further complicated by substantial heterogeneity in patient samples and outcome measures and subsequent substantial risk of bias across studies. Second, the variability across neuroimaging studies did not allow a meta-analytical approach, hence we interpreted this literature using a systematic review. Third, in the behavioral, cognitive and neuropathological meta-analyses, we combined comparable yet distinct study outcome measures such as different neuropsychological tests for memory and executive functions, questionnaires for neuropsychiatric/behavioral features, or staining methods and selection of brain regions for histopathological assessment of amyloid-β and tau. Fourth, we did not account for possible co-pathologies (e.g., Lewy bodies) that may contribute to the clinical phenotype. Fifth, there were only limited data on behavioral presentations of AD in diverse populations.

### Future directions

Akin to the development of diagnostic criteria for PCA, we consider the currently proposed provisional research criteria as a steppingstone towards internationally established consensus criteria for bvAD. For PCA, provisional research criteria were first proposed by two research groups and were subsequently applied by other groups to establish a PCA diagnosis for several years^46,47^, followed by widely supported formal diagnostic criteria based on consensus by an international working group.^10^ Similarly, our bvAD criteria should improve the consistency and reliability of future research and possibly aid in the clinical assessment of bvAD, which in turn would enhance the diagnostic accuracy of future bvAD criteria to be established by a working group of worldwide experts. There are several promising novel biomarkers and behavioral features that could be included in future bvAD criteria, for example more objective measurements of behavior like social cognition in conjunction with biometric information (e.g., eye-tracking, face reading, galvanic skin response)^48^ or blood-based biomarkers of AD pathology (e.g., p-tau, amyloid-β) and neurodegeneration (e.g., neurofilament light chain [NfL]).^49^ Furthermore, the diagnostic utility of potential bvAD-specific features (e.g., relatively preserved disease insight, presence of hallucinations and delusions) or measures of disease severity (e.g., the FTLD-modified Clinical Dementia Rating scale^50^) should be further investigated.

## CONCLUSION

Although the existence of bvAD is acknowledged in the most recent diagnostic/research criteria for AD dementia^8,9^, there currently does not exist a set of criteria that provide specific recommendations for the diagnosis of bvAD. Our systematic review and meta-analyses of the current bvAD literature indicate that bvAD is clinically most similar to bvFTD, while it shares most pathophysiological features with tAD. Based on these insights, we here provide the first provisional research criteria for bvAD aimed at improving the consistency and reliability of future research, and potentially facilitating clinical assessment of bvAD.

## Data Availability

Data are available upon reasonable request.

## Acknowledgements

The authors would like to thank Drs. Jeffrey Phillips, Dennis Irwin, David Bergeron and Claire Boutoleau-Bretonnière for providing missing data from their manuscripts that could now be used in the meta-analyses presented in this manuscript. Furthermore, we would like to thank Prof. Oskar Hansson and Dr. Sebastian Palmqvist for their advice on how to incorporate biofluid vs neuroimaging biomarkers into the research criteria for bvAD.

## Funding

This project has received funding from the Netherlands Organization for Health Research and Development, ZonMw (70-73305-98-1214 to Rik Ossenkoppele [PI] and Janne Papma [PI]). Research of the Alzheimer Center Amsterdam is part of the neurodegeneration research programme of Amsterdam Neuroscience. The Alzheimer Center Amsterdam is supported by Stichting Alzheimer Nederland and Stichting VUmc fonds. Research at UCSF is supported by the following grants: P30-AG062422 (BLM, GDR), P01-AG019724 (BLM, GDR), R35 AG072362 (GDR), R01-AG038791 (GDR), R01-NS050915 (MLGT), R01 AG045611 (GDR), Rainwater Charitable Foundation (GDR). Research at CHU de Québec is supported by the Chaire de recherche sur les aphasies primaires progressives – Fondation de la famille Lemaire (RJL).

## Conflicts of interest/Competing interests

GDR receives research support from NIH, Alzheimer Association, American College of Radiology, Avid Radiopharmaceuticals, GE Healthcare, Life Molecular Imaging. In the past 2 years he has received consulting fees from Axon Neurosciences, Eisai, GE Healthcare, Johnson & Johnson, Merck. He is an Associate Editor for JAMA Neurology.

The other authors report no conflict of interest.

## Notes

### Competing Interest Statement

The authors have declared no competing interest.

### Author Declarations

All work presented has been approved by local IRB and/or medical ethics review boards.

## REFERENCES

1. Scheltens P, De Strooper B, Kivipelto M, et al. Alzheimer’s disease. Lancet. 2021;397(10284):1577–1590.

2. Graff-Radford J, Yong KXX, Apostolova LG, et al. New insights into atypical Alzheimer’s disease in the era of biomarkers. Lancet Neurol. 2021;20(3):222–234.

3. Rabinovici GD, Rosen HJ, Alkalay A, et al. Amyloid vs FDG-PET in the differential diagnosis of AD and FTLD. Neurology. 2011;77(23):2034–2042.

4. Ossenkoppele R, Prins ND, Pijnenburg YA, et al. Impact of molecular imaging on the diagnostic process in a memory clinic. Alzheimers Dement. 2013;9(4):414–421.

5. Forman MS, Farmer J, Johnson JK, et al. Frontotemporal dementia: Clinicopathological correlations. Annals of Neurology. 2006;59(6):952–962.

6. Beach TG, Monsell SE, Phillips LE, Kukull W. Accuracy of the clinical diagnosis of Alzheimer disease at National Institute on Aging Alzheimer Disease Centers, 2005-2010. J Neuropathol Exp Neurol. 2012;71(4):266–273.

7. Cummings J, Aisen PS, Apostolova LG, Atri A, Salloway S, Weiner M. Aducanumab: Appropriate Use Recommendations. The Journal of Prevention of Alzheimer’s Disease 2021.

8. Dubois B, Villain N, Frisoni GB, et al. Clinical diagnosis of Alzheimer’s disease: recommendations of the International Working Group. Lancet Neurol. 2021;20(6):484–496.

9. Jack CR, Jr., Bennett DA, Blennow K, et al. NIA-AA Research Framework: Toward a biological definition of Alzheimer’s disease. Alzheimers Dement. 2018;14(4):535–562.

10. Crutch SJ, Schott JM, Rabinovici GD, et al. Consensus classification of posterior cortical atrophy. Alzheimers Dement. 2017;13(8):870–884.

11. Gorno-Tempini ML, Hillis AE, Weintraub S, et al. Classification of primary progressive aphasia and its variants. Neurology. 2011;76(11):1006–1014.

12. Townley RA, Graff-Radford J, Mantyh WG, et al. Progressive dysexecutive syndrome due to Alzheimer’s disease: a description of 55 cases and comparison to other phenotypes. Brain Commun. 2020;2(1):fcaa068.

13. Rascovsky K, Hodges JR, Knopman D, et al. Sensitivity of revised diagnostic criteria for the behavioural variant of frontotemporal dementia. Brain. 2011;134(Pt 9):2456–2477.

14. Cummings J. The Neuropsychiatric Inventory: Development and Applications. Journal of geriatric psychiatry and neurology. 2020;33(2):73–84.

15. Bergeron D, Sellami L, Poulin S, Verret L, Bouchard RW, Laforce R. The Behavioral/Dysexecutive Variant of Alzheimer’s Disease: A Case Series with Clinical, Neuropsychological, and FDG-PET Characterization. Dementia and Geriatric Cognitive Disorders. 2021;49(5):518–525.

16. Lehingue E, Gueniat J, Jourdaa S, et al. Improving the Diagnosis of the Frontal Variant of Alzheimer’s Disease with the DAPHNE Scale. Journal of Alzheimers Disease. 2021;79(4):1735–1745.

17. Phillips JS, Da Re F, Dratch L, et al. Neocortical origin and progression of gray matter atrophy in nonamnestic Alzheimer’s disease. Neurobiol Aging. 2018;63:75–87.

18. Eikelboom WS, van den Berg E, Singleton EH, et al. Neuropsychiatric and Cognitive Symptoms Across the Alzheimer Disease Clinical Spectrum: Cross-sectional and Longitudinal Associations. Neurology. 2021.

19. Blennerhassett R, Lillo P, Halliday GM, Hodges JR, Kril JJ. Distribution of pathology in frontal variant Alzheimer’s disease. Journal of Alzheimer’s disease: JAD. 2014;39(1):63–70.

20. de Souza LC, Bertoux M, Funkiewiez A, et al. Frontal presentation of Alzheimer’s disease: a series of patients with biological evidence by CSF biomarkers. Dementia & neuropsychologia. 2013;7(1):66–74.

21. Léger GC, Banks SJ. Neuropsychiatric symptom profile differs based on pathology in patients with clinically diagnosed behavioral variant frontotemporal dementia. Dementia and geriatric cognitive disorders. 2014;37(1-2):104-112.

22. Mendez MF, Joshi A, Tassniyom K, Teng E, Shapira JS. Clinicopathologic differences among patients with behavioral variant frontotemporal dementia. Neurology. 2013;80(6):561–568.

23. Ossenkoppele R, Pijnenburg YA, Perry DC, et al. The behavioural/dysexecutive variant of Alzheimer’s disease: clinical, neuroimaging and pathological features. Brain. 2015;138(Pt 9):2732–2749.

24. Perry DC, Brown JA, Possin KL, et al. Clinicopathological correlations in behavioural variant frontotemporal dementia. Brain. 2017;140(12):3329–3345.

25. Phillips JS, Da Re F, Irwin DJ, et al. Longitudinal progression of grey matter atrophy in non-amnestic Alzheimer’s disease. Brain. 2019;142(6):1701–1722.

26. Therriault J, Pascoal TA, Savard M, et al. Topographical distribution of amyloid-β, tau and atrophy in behavioral / dysexecutive AD patients. Neurology. 2020:10.1212/WNL.0000000000011081.

27. Singleton EH, Pijnenburg YAL, Sudre CH, et al. Investigating the clinico-anatomical dissociation in the behavioral variant of Alzheimer disease. Alzheimers Res Ther. 2020;12(1):148.

28. Bergeron D, Beauregard JM, Jean G, et al. Posterior Cingulate Cortex Hypometabolism in Non-Amnestic Variants of Alzheimer’s Disease. Journal of Alzheimers Disease. 2020;77(4):1569–1577.

29. Sala A, Caprioglio C, Santangelo R, et al. Brain metabolic signatures across the Alzheimer’s disease spectrum. Eur J Nucl Med Mol Imaging. 2020;47(2):256–269.

30. Woodward MC, Rowe CC, Jones G, Villemagne VL, Varos TA. Differentiating the frontal presentation of Alzheimer’s disease with FDG-PET. Journal of Alzheimer’s disease: JAD. 2015;44(1):233–242.

31. Wang Y, Shi Z, Zhang N, et al. Spatial Patterns of Hypometabolism and Amyloid Deposition in Variants of Alzheimer’s Disease Corresponding to Brain Networks: a Prospective Cohort Study. Molecular imaging and biology. 2019;21(1):140–148.

32. Singleton E, Hansson O, Pijnenburg YAL, et al. Heterogeneous distribution of tau pathology in the behavioural variant of Alzheimer’s disease. Journal of Neurology, Neurosurgery & Psychiatry. 2021:jnnp-2020-325497.

33. Johnson JK, Diehl J, Mendez MF, et al. Frontotemporal lobar degeneration: demographic characteristics of 353 patients. Archives of neurology. 2005;62(6):925–930.

34. Mattsson N, Groot C, Jansen WJ, et al. Prevalence of the apolipoprotein E epsilon4 allele in amyloid beta positive subjects across the spectrum of Alzheimer’s disease. Alzheimers Dement. 2018;14(7):913–924.

35. Seeley WW, Menon V, Schatzberg AF, et al. Dissociable intrinsic connectivity networks for salience processing and executive control. J Neurosci. 2007;27(9):2349–2356.

36. Snowden JS, Stopford CL, Julien CL, et al. Cognitive phenotypes in Alzheimer’s disease and genetic risk. Cortex; a journal devoted to the study of the nervous system and behavior. 2007;43(7):835–845.

37. Woodward M, Jacova C, Black SE, et al. Differentiating the frontal variant of Alzheimer’s disease. International Journal of Geriatric Psychiatry. 2010;25(7):732–738.

38. Ossenkoppele R, Jansen WJ, Rabinovici GD, et al. Prevalence of amyloid PET positivity in dementia syndromes: a meta-analysis. JAMA. 2015;313(19):1939–1949.

39. Rabinovici GD, Furst AJ, Neil JP, et al. 11C-PIB PET imaging in Alzheimer disease and frontotemporal lobar degeneration. Neurology. 2007;68(15):1205.

40. Ashton NJ, Janelidze S, Al Khleifat A, et al. A multicentre validation study of the diagnostic value of plasma neurofilament light. Nat Commun. 2021;12(1):3400.

41. Leuzy A, Ashton NJ, Mattsson-Carlgren N, et al. 2020 update on the clinical validity of cerebrospinal fluid amyloid, tau, and phospho-tau as biomarkers for Alzheimer’s disease in the context of a structured 5-phase development framework. Eur J Nucl Med Mol Imaging. 2021.

42. Ossenkoppele R, Rabinovici GD, Smith R, et al. Discriminative Accuracy of [18F]flortaucipir Positron Emission Tomography for Alzheimer Disease vs Other Neurodegenerative Disorders. JAMA. 2018;320(11):1151–1162.

43. Bischof GN, Dodich A, Boccardi M, et al. Clinical validity of second-generation tau PET tracers as biomarkers for Alzheimer’s disease in the context of a structured 5-phase development framework. Eur J Nucl Med Mol Imaging. 2021;48(7):2110–2120.

44. Jack CR, Wiste HJ, Botha H, et al. The bivariate distribution of amyloid-beta and tau: relationship with established neurocognitive clinical syndromes. Brain. 2019;142(10):3230–3242.

45. Fleisher AS, Pontecorvo MJ, Devous MD, Sr., et al. Positron Emission Tomography Imaging With [18F]flortaucipir and Postmortem Assessment of Alzheimer Disease Neuropathologic Changes. JAMA Neurol. 2020;77(7):829–839.

46. Mendez MF, Ghajarania M, Perryman KM. Posterior cortical atrophy: clinical characteristics and differences compared to Alzheimer’s disease. Dementia and geriatric cognitive disorders. 2002;14(1):33–40.

47. Tang-Wai DF, Graff-Radford NR, Boeve BF, et al. Clinical, genetic, and neuropathologic characteristics of posterior cortical atrophy. Neurology. 2004;63(7):1168–1174.

48. Adolphs R. Conceptual challenges and directions for social neuroscience. Neuron. 2010;65(6):752–767.

49. Hansson O. Biomarkers for neurodegenerative diseases. Nat Med. 2021;27(6):954–963.

50. Borroni B, Agosti C, Premi E, et al. The FTLD-modified Clinical Dementia Rating scale is a reliable tool for defining disease severity in frontotemporal lobar degeneration: evidence from a brain SPECT study. Eur J Neurol. 2010;17(5):703–707.

