## Supplement for "Provisional research criteria for the behavioral variant of Alzheimer’s disease A systematic review and meta-analysis"

### SUPPLEMENTAL DATA

**Table of content**


| Table/Figure | Title | Page |
|---|---|---|
| Table S1 | PRISMA guidelines | 2-5 |
| Table S2 | Full database queries | 6 |
| Table S3 | Risk of bias assessment | 7 |
| Table S4 | Characteristics of included studies | 8-16 |
| Table S5 | Percentage of bvFTD features and NPI items in bvAD, bvFTD and tAD | 17 |
| Table S6 | Functional connectivity and white matter hyperintensities in bvAD | 18 |
| Figure S1 | Funnel plots for behavorial/neuropsychiatric data in meta-analysis | 19 |
| Figure S2 | Funnel plots for cognitive data in meta-analysis | 20 |
| Figure S3 | Funnel plots for neuropathological data in meta-analysis | 21 |
| Figure S4 | Risk of bias assessment summary | 22 |
| Figure S5 | Flow chart of study inclusion | 23 |
| Figure S6 | Results of meta-analysis for behavorial and neuropsychiatric seperately | 24 |
| Figure S7 | Meta-analyses for neuropathological data in bvAD vs typical AD | 25 |
| References | Reference list Supplement | 26-30 |

**Table S1.** PRISMA guidelines

| **Section and Topic** | **Item #** | **Checklist item** | **Location where item is reported** |
|---|---|---|---|
| **TITLE** | | | |
| Title | 1 | Identify the report as a systematic review. | 1 |
| **ABSTRACT** | | | |
| Abstract | 2 | See the PRISMA 2020 for Abstract checklist. | 2-4 |
| **INTRODUCTION** | | | |
| Rationale | 3 | Describe the rationale for the review in the context of existing knowledge. | 5 |
| Objectives | 4 | Provide an explicit statement of the objective(s) or question(s) the review addresses. | 5 |
| **METHODS** | | | |
| Eligibility criteria | 5 | Specify the inclusion and exclusion criteria for the review and how studies were grouped for the syntheses. | 6 |
| Information sources | 6 | Specify all databases, registers, websites, organisations, reference lists and other sources searched or consulted to identify studies. Specify the date when each source was last searched or consulted. | 6 |
| Search strategy | 7 | Present the full search strategies for all databases, registers and websites, including any filters and limits used. | 6& supplemental data |
| Selection process | 8 | Specify the methods used to decide whether a study met the inclusion criteria of the review, including how many reviewers screened each record and each report retrieved, whether they worked independently, and if applicable, details of automation tools used in the process. | 6&7 |
| Data collection process | 9 | Specify the methods used to collect data from reports, including how many reviewers collected data from each report, whether they worked independently, any processes for obtaining or confirming data from study investigators, and if applicable, details of automation tools used in the process. | 6&7 |
| Data items | 10a | List and define all outcomes for which data were sought. Specify whether all results that were compatible with each outcome domain in each study were sought (e.g. for all measures, time points, analyses), and if not, the methods used to decide which results to collect. | 6&7 |

| Section and Topic | Item # | Checklist item | Location where item is reported |
|---|---|---|---|
| | 10b | List and define all other variables for which data were sought (e.g. participant and intervention characteristics, funding sources). Describe any assumptions made about any missing or unclear information. | 6&7 |
| Study risk of bias assessment | 11 | Specify the methods used to assess risk of bias in the included studies, including details of the tool(s) used, how many reviewers assessed each study and whether they worked independently, and if applicable, details of automation tools used in the process. | 7 |
| Effect measures | 12 | Specify for each outcome the effect measure(s) (e.g. risk ratio, mean difference) used in the synthesis or presentation of results. | 7 |
| Synthesis methods | 13a | Describe the processes used to decide which studies were eligible for each synthesis (e.g. tabulating the study intervention characteristics and comparing against the planned groups for each synthesis (item #5)). | 6 |
| | 13b | Describe any methods required to prepare the data for presentation or synthesis, such as handling of missing summary statistics, or data conversions. | 6 |
| | 13c | Describe any methods used to tabulate or visually display results of individual studies and syntheses. | 7 |
| | 13d | Describe any methods used to synthesize results and provide a rationale for the choice(s). If meta-analysis was performed, describe the model(s), method(s) to identify the presence and extent of statistical heterogeneity, and software package(s) used. | 7 |
| | 13e | Describe any methods used to explore possible causes of heterogeneity among study results (e.g. subgroup analysis, meta-regression). | n/a |
| | 13f | Describe any sensitivity analyses conducted to assess robustness of the synthesized results. | n/a |
| Reporting bias assessment | 14 | Describe any methods used to assess risk of bias due to missing results in a synthesis (arising from reporting biases). | 7 |
| Certainty assessment | 15 | Describe any methods used to assess certainty (or confidence) in the body of evidence for an outcome. | 7 |
| **RESULTS** | | | |
| Study selection | 16a | Describe the results of the search and selection process, from the number of records identified in the search to the number of studies included in the review, ideally using a flow diagram. | 8 & supplemental data |

| Section and Topic | Item # | Checklist item | Location where item is reported |
|---|---|---|---|
| | 16b | Cite studies that might appear to meet the inclusion criteria, but which were excluded, and explain why they were excluded. | n/a |
| Study characteristics | 17 | Cite each included study and present its characteristics. | Supplemental data |
| Risk of bias in studies | 18 | Present assessments of risk of bias for each included study. | Supplemental data |
| Results of individual studies | 19 | For all outcomes, present, for each study: (a) summary statistics for each group (where appropriate) and (b) an effect estimate and its precision (e.g. confidence/credible interval), ideally using structured tables or plots. | 8&9 & Figure 1&2 |
| Results of syntheses | 20a | For each synthesis, briefly summarise the characteristics and risk of bias among contributing studies. | 9 |
| | 20b | Present results of all statistical syntheses conducted. If meta-analysis was done, present for each the summary estimate and its precision (e.g. confidence/credible interval) and measures of statistical heterogeneity. If comparing groups, describe the direction of the effect. | 8-10 |
| | 20c | Present results of all investigations of possible causes of heterogeneity among study results. | n/a |
| | 20d | Present results of all sensitivity analyses conducted to assess the robustness of the synthesized results. | n/a |
| Reporting biases | 21 | Present assessments of risk of bias due to missing results (arising from reporting biases) for each synthesis assessed. | 7 & Supplemental data |
| Certainty of evidence | 22 | Present assessments of certainty (or confidence) in the body of evidence for each outcome assessed. | 8-10 |
| **DISCUSSION** | | | |
| Discussion | 23a | Provide a general interpretation of the results in the context of other evidence. | 10-14 |
| | 23b | Discuss any limitations of the evidence included in the review. | 14 |
| | 23c | Discuss any limitations of the review processes used. | 14 |
| | 23d | Discuss implications of the results for practice, policy, and future research. | 15&16 |

| Section and Topic | Item # | Checklist item | Location where item is reported |
|---|---|---|---|
| **OTHER INFORMATION** | | | |
| Registration and protocol | 24a | Provide registration information for the review, including register name and registration number, or state that the review was not registered. | 6 |
| | 24b | Indicate where the review protocol can be accessed, or state that a protocol was not prepared. | 6 |
| | 24c | Describe and explain any amendments to information provided at registration or in the protocol. | n/a |
| Support | 25 | Describe sources of financial or non-financial support for the review, and the role of the funders or sponsors in the review. | 13 |
| Competing interests | 26 | Declare any competing interests of review authors. | 13 |
| Availability of data, code and other materials | 27 | Report which of the following are publicly available and where they can be found: template data collection forms; data extracted from included studies; data used for all analyses; analytic code; any other materials used in the review. | 7 |

**Table S2.** Full database queries used in the present study

| Database | Search no. | Search terms | No. of studies |
|---|---|---|---|
| PubMed/medline | 1 | (Alzheimer*[Title]) AND (behavio* variant[Title] OR executive variant[Title] OR dysexecutive variant[Title] OR behavio*/dysexecutive AD[Title] OR frontal variant[Title] OR frontal presentation[Title] OR nonamnestic[Title] OR non-amnestic[Title] OR heterogene*[Title] OR atypical[Title]) | 492 |
| | 2 | (frontotemporal dementia[Title]) AND (pathology[Title] OR clinicopathologic* [Title]) | 73 |
| Web of Science | 1 | TITLE: (Alzheimer*) AND TITLE: (behavio* variant OR executive variant OR dysexecutive variant OR behavio*/dysexecutive AD OR frontal variant OR frontal presentation OR nonamnestic OR non-amnestic OR heterogene* OR atypical) | 581 |
| | 2 | TITLE: (frontotemporal dementia) AND TITLE: (pathology OR clinicopathologic*) | 111 |

**Table S3.** Risk of bias assessment per domain according to the ROBINS-I tool per study included in the meta-analyses.

| Study | D1 Bias due to confounding | D2 Bias in selection of participants into the study | D3 Bias in classification of interventions | D4 Bias due to deviations from intended interventions | D5 Bias due to missing data | D6 Bias in measurement of outcomes | D7 Bias in selection of the reported result | Overal bias |
|---|---|---|---|---|---|---|---|---|
| Woodward et al. 2010[1] | Y | Y | NA | NA | PY | N | N | Serious risk of bias |
| Balasa et al. 2011[2] | PY | PY | NA | NA | N | N | N | Moderate risk of bias |
| de Souza et al. 2013[3] | Y | PN | NA | NA | N | N | N | Moderate risk of bias |
| Mendez et al. 2013[4] | Y | PN | NA | NA | N | N | N | Moderate risk of bias |
| Fernández-Calvo et al. 2013[5] | PY | PY | NA | NA | PY | N | N | Moderate risk of bias |
| Blennerhassett et al. 2014[6] | PY | PN | NA | NA | PN | N | N | Moderate risk of bias |
| Ossenkoppele et al. 2015[7] | PN | PN | NA | NA | PN | N | N | Moderate risk of bias |
| Phillips et al. 2018[8] | PN | PY | NA | NA | PN | PY | N | Moderate risk of bias |
| Sala et al. 2020[9] | Y | PN | NA | NA | PY | N | PY | Moderate risk of bias |
| Therriault et al. 2020[10] | PN | PY | NA | NA | PY | N | N | Moderate risk of bias |
| Bergeron et al. 2020[11] | PY | Y | NA | NA | PY | PN | PY | Moderate risk of bias |
| Singleton et al. 2021[12] | PN | PN | NA | NA | PN | PY | N | Moderate risk of bias |
| Lehingue et al. 2021[13] | PY | Y | NA | NA | PY | PN | N | Serious risk of bias |

*Y*=yes, *N*=no, *PY*=possible yes, *PN*=possible no.

**Table S4.** Characteristics of included studies in chronological order

| Study | Design | Country | N | Participants | Controls | Age | Sex | MMSE | Confirmation of AD | Main topic of group study | Type of data in case studies | | | | | |
|---|---|---|---|---|---|---|---|---|---|---|---|---|---|---|---|---|
| | | | | | | | | | | | CL | CO | SO | NI | PA | GE |
| Brun et al. 1976[14] | Case study | Sweden | 5 | bvAD | - | 56 (5.69) | 60 | n/a | Autopsy | | X | | | | X | |
| Shibayama et al. 1978[15] | Case study | Japan | 1 | bvAD | - | 70 | 0 | n/a | Autopsy | | X | | | | X | |
| Shuttleworth 1984[16] | Case study | US | 2 | bvAD | - | 49 (3) | 0 | n/a | No | | X | X | | X | | |
| Brun 1987[17] | Case study | Sweden | 2 | bvAD | - | 75 (6) | 100 | n/a | Autopsy | | X | | | | X | |
| Perani et al. 1988[18] | Case study (within a cross-sectional observational study) | Italy | 1 | bvAD | - | 56 | 100 | n/a | No | | X | X | | X | | |
| Bird et al. 1989[19] | Cross-sectional observational study | US | 2 | bvAD | - | 66 (1) | 0 | n/a | Autopsy | | X | | | | X | X |
| Grady et al. 1990[20] | Cross-sectional observational study | US | | bvAD | Subgroups of AD | 71.5 | 20 | 8 (7) | No | Neuroimaging | | | | | | |
| Molchan et al. 1990[21] | Case study | US | 2 | bvAD | - | 58 (3) | 50 | 12.5 (0.5) | No | | X | X | | | | |

| | | | | | | | | | | | | | | | | |
|---|---|---|---|---|---|---|---|---|---|---|---|---|---|---|---|---|
| Raux et al. 2000[22] | Case series | France | 3 | bvAD | - | 49.3 (10.4) | 100 | n/a | Genetic | | | | | | | |
| Rippon et al. 2003[23] | Case study | US | 2 | bvAD | - | 43.5 (1.5) | 0 | 18.5 (5.5) | | | X | X | | X | X | X |
| Yokota et al. 2003[24] | Case study | Japan | 3 | bvAD | - | 33.7 (4.5) | 66.7 | n/a | Autopsy | | X | | | | X | X |
| Doran & Larner 2004[25] | Case study | | 2 | bvAD | - | 49 (0) | 50 | n/a | Genetic | | X | | | | X | X |
| Kertesz et al. 2005[26] | Cross-sectional observational cohort study | Canada | 1 | bvAD | - | n/a | n/a | n/a | Autopsy | Clinicopatholo gical | | | | | | |
| Shi et al. 2005[27] | Cross-sectional cohort study | China | 1 | bvAD | - | 59 | 0 | n/a | Autopsy | Clinicopatholo gical | | | | | | |
| Forman et al. 2006[28] | Cohorts study | US | 19 | bvAD | FTLD | 60.3 | 47 | 20.1 (2-29) | Autopsy | Clinicopatholo gical | | | | | | |
| Larner 2006[29] | Case study | UK | 2 | bvAD | - | 54 (2) | 0 | 16.5 (0.5) | No | | X | X | | X | | X |
| Alladi et al. 2007[30] | Corss-sectional observational cohort study | UK | 2 | bvAD | atypical AD | n/a | n/a | n/a | Autopsy | Clinicopatholo gical | | | | | | |
| Rabinovici et al. 2007[31] | Cross-sectional observational study | US | 2 | bvAD | - | 54 (1) | 50 | 22.5 2.5 | Amyloid PET | Neuroimaging | | | | | | |
| Snowden et al. 2007[32] | Cross-sectional observational cohort study | UK | 12 | bvAD | AD & atypical AD | 49 (8) | 25 | n/a | No | Cognitive & Genetic | | | | | | |

| | | | | | | | | | | | | | | | | |
|---|---|---|---|---|---|---|---|---|---|---|---|---|---|---|---|---|
| Taylor et al. 2008[33] | Case study | UK | 1 | bvAD | - | 66 | 0 | 28 | Autopsy | | X | X | | X | X | |
| Kile et al. 2009[34] | Case study | US | 1 | bvAD | - | n/a | 0 | 30 | Autopsy | | | | | | | |
| Bigio et al. 2010[35] | Cross-sectional observational study | US | 10 | bvAD | AD & FTD | 58 (6.5) | 30 | n/a | Autopsy | | X | | | | X | |
| Habek et al. 2010[36] | Case study | Croatia | 1 | bvAD | - | 56 | 0 | n/a | Biopsy | | X | | | X | | |
| Lehman et al. 2010[37] | Cross-sectional observational study | UK | 2 | bvAD | AD & FTD | 59 (1.4) | 50 | 9.5 (0.7) | Autopsy | Clinicopatholo gical & Neuroimaging | | | | | | |
| Piscopo et al. 2010[38] | Case study | Italy | 1 | bvAD | - | 63 | n/a | 11 | Genetic | | X | X | | | | X |
| Woodwa rd et al. 2010[1] | Cross-sectional observational cohort study | Canada | 18 | bvAD | AD & FTD | 74.7 (7) | 44.4 | 18.6 5.9 | No | Clinical & Genetic | | | | | | |
| Balasa et al. 2011[2] | Cross-sectional observational cohort study | Spain | 7 | bvAD | AD & atypical AD | 55.6 (3.7) | 28.6 | n/a | Autopsy | Clinicopatholo gical & Genetic | | | | | | |
| Rabinovi ci et al. 2011[39] | | US | 3 | bvAD | AD, FTD & CN | n/a | n/a | n/a | PET | Neuroimaging | | | | | | |
| Snowden et al. 2011[40] | Cross-sectional observational study | UK | 2 | bvAD | - | 60.5 (6.5) | 50 | n/a | Autopsy | Clinicopatholo gical | | | | | | |

| | | | | | | | | | | | | | | | |
|---|---|---|---|---|---|---|---|---|---|---|---|---|---|---|---|
| Whitwell et al. 2011[41] | Cross-sectional observational study | US | 3 | bvAD | AD & CN | 58.33 (3.3) | 33.3 | n/a | Autopsy | Neuroimaging | | | | | |
| Borroni et al. 2012[42] | Case study | Italy | 1 | bvAD | - | 68 | 0 | 21 | Genetic & CSF | | | | | | |
| Duker et al. 2012[43] | Case study | US | 1 | bvAD | - | 58 | 0 | n/a | No | | X | X | | X | |
| Wallon et al. 2012[44] | Case series | France | 8 | bvAD | - | n/a | n/a | n/a | Genetic | Genetic | | | | | |
| De Souza et al. 2013[3] | Case series | France | 8 | bvAD | AD, bvFTD & CN | 63.5 (8.9) | 12.5 | 17.6 5.6 | CSF | Cognitive & Neuroimaging | | | | | |
| Fernandez-Calvo et al. 2013[5] | Cross-sectional observational study | Spain | 13 | bvAD | AD & CN | 72.8 (7.6) | 31 | 22.5 2.1 | No | Cognitive & Neuropsychiatric | | | | | |
| Herrero-San Martin et al. 2013[45] | Case study | Spain | 2 | bvAD | - | 56 (4) | 50 | n/a | Autopsy | | X | | | | X |
| Marini et al. 2013[46] | Case study | Italy | 1 | bvAD | - | 59 | 100 | n/a | Genetic | | | | | | |
| Mendez et al. 2013[46] | Cross-sectional observational cohort study | US | 21 | bvAD | FTLD | 69.3 (8.3) | 14.3 | 13.3 9.4 | Autopsy | Clinicopathological | | | | | |

| | | | | | | | | | | | | | | |
|---|---|---|---|---|---|---|---|---|---|---|---|---|---|---|
| Blennerhassett et al. 2014[6] | Cross-sectional observational study | Australia | 6 | bvAD | AD & bvFTD | 68 (14) | 33.4 | n/a | Autopsy | Pathological | | | | |
| Leger et al. 2014[47] | Cross-sectional observational study | US | 31 | bvAD | FTLD | n/a | n/a | n/a | Autopsy | Clinicopathological | | | | |
| Nijgaard et al. 2014[48] | Case study | US | 1 | bvAD | - | 52 | 0 | 30 | Genetic | | X | X | X | X |
| Balasa et al. 2015[49] | Cross-sectional observational study | Spain | 13 | bvAD | FTLD | n/a | n/a | n/a | Autopsy | Clinicopathological | | | | |
| Ossenkoppele et al. 2015[7] | Cross-sectional observational study | Netherlands & US | 55 | bvAD | AD, bvFTD & CN | 64.7 (8.8) | 27.3 | 22.5 5.4 | Autopsy | Clinical, Cognitive & Neuroimaging | | | | |
| Paterson et al. 2015[50] | Cross-sectional observational study | UK | 8 | bvAD | AD, atypical AD & CN | 61.5 (6.4) | 62.5 | 17.4 6.1 | CSF/PET/ autopsy | Cerebrospinal fluid | | | | |
| Woodward et al. 2015[51] | Cross-sectional observational study | NA | 13 | bvAD | AD | 81.6 (4.1) | 38.5 | 23.9 | No | Neuroimaging | | | | |
| Li et al. 2016[52] | Case study | China | 1 | bvAD | bvFTD | 71 | 100 | 15 | PET | | X | X | X | X |
| Ossenkoppele et al. 2016[53] | Cross-sectional observational study | US | 1 | bvAD | AD, atypical AD & CN | 59 | 0 | 21 | PET | | | | | |
| Scialo et al. 2016[54] | Case study | Italy | 1 | bvAD | - | 68 | 100 | 27 | PET | | X | X | X | X |

| | | | | | | | | | | | | | | | | |
|---|---|---|---|---|---|---|---|---|---|---|---|---|---|---|---|---|
| Dickerson et al. 2017[55] | Case series | US | 1 | bvAD | - | 62 | 100 | n/a | CSF | | X | X | | X | | |
| Duclos et al. 2017[56] | Case study | France | 1 | bvAD | CN | 60 | 100 | n/a | CSF | | X | X | X | X | | |
| Kawakatsu et al. 2017[57] | Case series | Japan | 3 | bvAD | - | 57.7 (1.3) | 33.3 | n/a | Autopsy | | X | | | X | X | |
| Oboudiyat et al. 2017[58] | Cross-sectional observational study | US | 2 | bvAD | - | n/a | n/a | n/a | CSF & autopsy | Cerebrospinal fluid | | | | | | |
| Perry et al. 2017[59] | Cross-sectional observational study | US | 15 | bvAD | FTLD | 62.8 (43-83) | 33.3 | 19.8 6.9 | Autopsy | Clinicopathological & Neuroimaging | | | | | | |
| Rawtaer et al. 2017[60] | Case study | Canada | 1 | bvAD | - | 68 | 0 | 11 | No | | X | X | | X | | X |
| Sawyer et al. 2017[61] | Case series | US | 3 | bvAD | - | 76.3 (3.1) | 33.3 | n/a | Autopsy | | X | X | | X | X | |
| Bagyinszky et al. 2018[62] | Case series | Korea | 1 | bvAD | - | 41 | 100 | 24 | Genetic | | X | X | | X | | X |
| Boon et al. 2018[63] | Cross-sectional observational study | Netherlands | 3 | bvAD | AD | 60.7 (1.3) | 0 | n/a | Autopsy | Pathological | | | | | | |
| Phillips et al. 2018[8] | Observational cross-sectional study | US | 22 | b/dAD | AD, atypical AD & CN | 64.3 (8.2) | 50 | 19.6 8.4 | CSF/autopsy | Neuroimaging & Pathological | | | | | | |

| | | | | | | | | | | | | | | | | |
|---|---|---|---|---|---|---|---|---|---|---|---|---|---|---|---|---|
| Seo et al. 2018[64] | Retrospective observational study | US | 23 | bvAD | - | n/a | n/a | n/a | Autopsy | Clinicopathological | | | | | | |
| Whitwell et al. 2018[65] | Cross-sectional observational study | US | 6 | b/dAD | AD & atypical AD | n/a | n/a | n/a | PET | Neuroimaging | | | | | | |
| De Souza et al. 2019[66] | Case study | Brazil | 1 | bvAD | - | 68 | 100 | 29 | CSF | | X | X | X | X | | |
| Foiani et al. 2019[67] | Cross-sectional observational study | UK | 2 | bvAD | FTLD | n/a | n/a | n/a | CSF | Cerebrospinal fluid | | | | | | |
| Monacelli et al. 2019[68] | Case study | Italy | 1 | bvAD | - | 60 | 100 | 25 | Genetic | | X | X | | X | | X |
| Nolan et al. 2019[69] | Cross-sectional observational study | US | 5 | bvAD | AD, atypical AD & CN | 66.2 (4.8) | 20 | n/a | Autopsy | Pathological | | | | | | |
| Pawlowski et al. 2019[70] | Cross-sectional observational study | Germany | 8 | bvAD | AD & FTD | n/a | n/a | n/a | CSF | Clinical & Cerebrospinal fluid | | | | | | |
| Phillips et al. 2019[71] | Cross-sectional & longitudinal observational study | US | 12 | bvAD | AD, atypical AD & CN | 63.9 (59.7-69.5) | 41.7 | 23 (17-26) | CSF/autopsy | Neuroimaging | | | | | | |
| Pillai et al. 2019[72] | Cross-sectional observational study | US | 4 | b/dAD | AD & atypical AD | n/a | n/a | n/a | CSF | Cerebrospinal fluid | | | | | | |

| | | | | | | | | | | | | | | | | |
|---|---|---|---|---|---|---|---|---|---|---|---|---|---|---|---|---|
| Tan et al. 2019[73] | Cross-sectional observational study | Australia | 9 | bvAD | AD | 65 (10) | 22 | n/a | Autopsy | Pathological | | | | | | |
| Wang et al. 2019[74] | Cross-sectional obersational study | China | 13 | b/dAD | AD, atypical AD & CN | 68 (3.4) | 69.2 | 17 (5.6) | PET | Neuroimaging | | | | | | |
| Wong et al. 2019[75] | Case study | Australia | 1 | bvAD | AD, bvFTD & CN | 57 | 0 | n/a | PET | | X | X | X | X | | |
| Bergeron et al. 2020[11] | Cross-sectional observational study | Canada | 8 | b/dAD | Atypical AD & FTD | 61.6 | n/a | 20.7 | CSF/PET | Neuroimaging | | | | | | |
| Cai et al. 2020[76] | Case study | China | 1 | bvAD | - | 52 | 100 | n/a | PET | | X | X | | X | | X |
| Cousins et al. 2020[77] | Cross-sectional observational study | US | 2 | bvAD | AD, atypical AD, FTD & CN | n/a | n/a | n/a | | Clinicopathological & Cerebrospinal fluid | | | | | | |
| Li et al. 2020[78] | Case study | Taiwan | 1 | bvAD | - | 66 | 0 | 10 | PET | | X | X | | X | | X |
| Paquin et al. 2020[79] | Case study | Canada | 1 | bvAD | - | 60 | 100 | 28 | Tau and amyloid PET | | X | X | | | X | |
| Sala et al. 2020[9] | Cross-sectional observational study | Italy | 15 | b/dAD | AD & atypical AD | 62.47 (5.7) | 33.3 | 16.5 (5.2) | CSF | Neuroimaging | | | | | | |
| Scarioni et al. 2020[80] | Cross-sectional observational study | Netherlands | 35 | bvAD | FTLD | n/a | n/a | n/a | Autopsy | Clinicopathological | | | | | | |
| Singleton et al. 2020[81] | Cross-sectional observational study | US | 29 | bvAD | AD, bvFTD & CN | 64.4 (9.4) | 41 | 22 (5.9) | CSF/PET/ autopsy | Neuroimaging | | | | | | |

| | | | | | | | | | | | | | | | | |
|---|---|---|---|---|---|---|---|---|---|---|---|---|---|---|---|---|
| Therriault et al. 2020[10] | Cross-sectional observational study | Canada | 15 | b/dAD | AD & CN | 65.93 (8.8) | 60 | 19.6 (5.3) | Tau & amyloid PET | Neuroimaging | | | | | | |
| Bergeron et al. 2021[82] | Case series | Canada | 8 | bvAD | AD & bvFTD | 59.5 (7.9) | 25 | 22.3 (5.9) | CSF/PET | Cognivite & Neuropsychiatric & Neuroimaging | | | | | | |
| Lehingue et al. 2021[13] | Cross-sectional prospective observational study | France | 20 | bvAD | AD & bvFTD | 71.5 (66-76) | 35 | 25 (21-26) | CSF | Cognitive & Neuropsychiatric & Neuroimaging | | | | | | |
| Singleton et al. 2021[12] | Cross-sectional observational study | Netherlands, Sweden & US | 7 & 8 | bvAD | AD | 69.1 (8.4) & 66.6 (6.0) | 14.3 & 50.0 | 21.7 (2.8) | CSF/PET and autopsy | Neuroimaging & Pathological | | | | | | |
| Zhu et al. 2021[83] | Case study | China | 1 | bvAD | - | 63 | 0 | 3 | CSF & PET | | X | X | | X | | X |

Numbers are depicted as mean (sd). *CL*=clinical, *COG*=cognition, *SOC*=social cognition, *NI*=neuroimaging, *PA*=pathological, *GEN*=genetic, *CSF*=cerebrospinal fluid, *PET*=positron emission tomography, AD=Alzheimer's disease, *bvAD*=behavioral variant of Alzheimer's disease, *bvFTD*=behavioral variant frontotemporal dementia.

**Table S5**. Weighted mean percentage of patients with separate behavioral and neuropsychiatric symptoms in bvAD and bvFTD.

| Diagnosis | bvAD | bvFTD | tAD | P-value of $\chi^2$-test bvAD vs bvFTD | P-value of $\chi^2$-test bvAD vs tAD |
|---|---|---|---|---|---|
| bvFTD criteria, n□ | 148† | 313* | | | |
| Disinhibition | 60.80 | 68.58 | | 0.10 | NA |
| Apathy | 68.80 | 77.37 | | 0.05 | NA |
| Loss of empathy | 54.64 | 53.64 | | 0.83 | NA |
| Compulsiveness | 45.00 | 68.50 | | <0.00001* | NA |
| Hyperorality | 35.89 | 64.11 | | <0.00001* | NA |
| NPI, n◊ | 52 | 156 | 1090▪ | | |
| Eating changes | 41.33 | 44.64 | 31.4 | 0.57 | 0.12 |
| Night-time behaviors | 39.60 | 40.73 | 20.0 | 0.94 | 0.0003* |
| Irritability | 50.81 | 42.15 | 42.9 | 0.33 | 0.32 |
| Euphoria | 16.62 | 27.09 | 6.0 | 0.16 | 0.005* |
| Anxiety | 54.15 | 43.10 | 31.6 | 0.17 | 0.001* |
| Depression | 34.19 | 35.10 | 32.1 | 0.93 | 0.78 |
| Agitation | 67.85 | 43.42 | 16.2 | 0.003* | <0.00001* |
| Hallucination | 28.23 | 9.00 | 4.6 | 0.0003* | <0.00001* |
| Delusions | 36.62 | 13.42 | 9.3 | 0.0003* | <0.00001* |
| Motor behavior | 50.38 | 57.10 | 18.9 | 0.38 | <0.00001* |

*bvAD*=behavioral variant of Alzheimer's disease, *bvFTD*=behavioral variant frontotemporal dementia, *tAD*=typical Alzheimer's disease.

†Based on estimates from 7 group studies(de Souza et al. 2013[3], Mendez et al. 2013[4], Blennerhassett et al. 2014[6], Ossenkoppele et al. 2015[7], Perry et al. 2017[59], Leger et al. 2014[47], Phillips et al. 2019[71])
* Based on estimates from 4 group studies (Mendez et al. 2013[4], Ossenkoppele et al. 2015[7], Perry et al. 2017[59], Leger et al. 2014[47])
□ Percentages are based on percentage per symptoms assessed by NPI, clinical evaluation or chart reviews from studies specified above.
◊ Percentages are based on percentage per symptoms assessed by NPI from two studies (Mendez et al. 2013[4], Leger et al. 2014[47]).
▪ Based on a cohort of Aβ-positive AD dementia patients from the Amsterdam dementia cohort (Eikelboom et al.[84]).

**Table S6**. Results of functional connectivity and white matter hyperintensities in bvAD

| Study | Subjects | Age | Sex | MMSE | AD confirmation | Contrasts | Modality | Findings |
|---|---|---|---|---|---|---|---|---|
| **Functional connectivity** | | | | | | | | |
| Wang et al. 2019[74] | 13 b/dAD | 68.0 (3.4) | 30.77 | 17.0 (5.6) | PiB PET | 38 typical AD, 20 CU | FDG-PET | The left executive control network showed the highest goodness-of-fit in both b/dvAD and tAD and no differences in PiB PET uptake in network templates was observed |
| Phillips et al. 2019[71] | 12 bvAD | 16.0 [13.5, 18.0] | 58.3 | 23.0 [17.0, 26.0] | CSF/autopsy | 17 typical AD | Diffusion MRI | Higher node degree predicted greater annualized grey matter volume loss in both bvAD and typical AD groups and bvAD showed a less negative slope of association between node degree and longitudinal atrophy than typical AD |
| Singleton et al. 2020[81] | 29 bvAD | 64.4 (9.4) | 59.0 | 22.0 (5.9) | CSF/PET/autopsy | 28 typical AD, 28 bvFTD, 34 CU | FDG-PET | The anterior default mode network showed highest goodness-of-fit in bvAD (tAD <bvAD=bvFTD), and significantly less metabolic connectivity of the posterior cingulate cortex to the (right) prefrontal cortex was observed in bvAD compared to tAD |
| **White matter hyperintensities** | | | | | | | | |
| Singleton et al. 2020[81] | 29 bvAD | 64.4 (9.4) | 59.0 | 22.0 (5.9) | CSF/PET/autopsy | 28 typical AD, 28 bvFTD, 34 CU | FLAIR-MRI | In comparison to tAD, bvAD patients showed lower juxtacortical left temporal and subcortical WMHV and higher right temporal juxtacortical WMHV |

*b/*dAD=behavioral/dysexecutive variant of Alzheimer's disease, *bvAD*=behavioral variant of Alzheimer's disease, *bvFTD*=behavioral variant frontotemporal dementia, *tAD*=typical Alzheimer's disease, *CU*=cognitively unimpaired individuals, *CSF*=cerebrospinal fluid, *PET*=positron emission tomography, *MRI*=magnetic resonance imaging.

**Figure S1**. Funnel plots of meta-analyses of behavorial/neuropsychiatric data for behavioral variant AD versus typical AD and bvFTD.

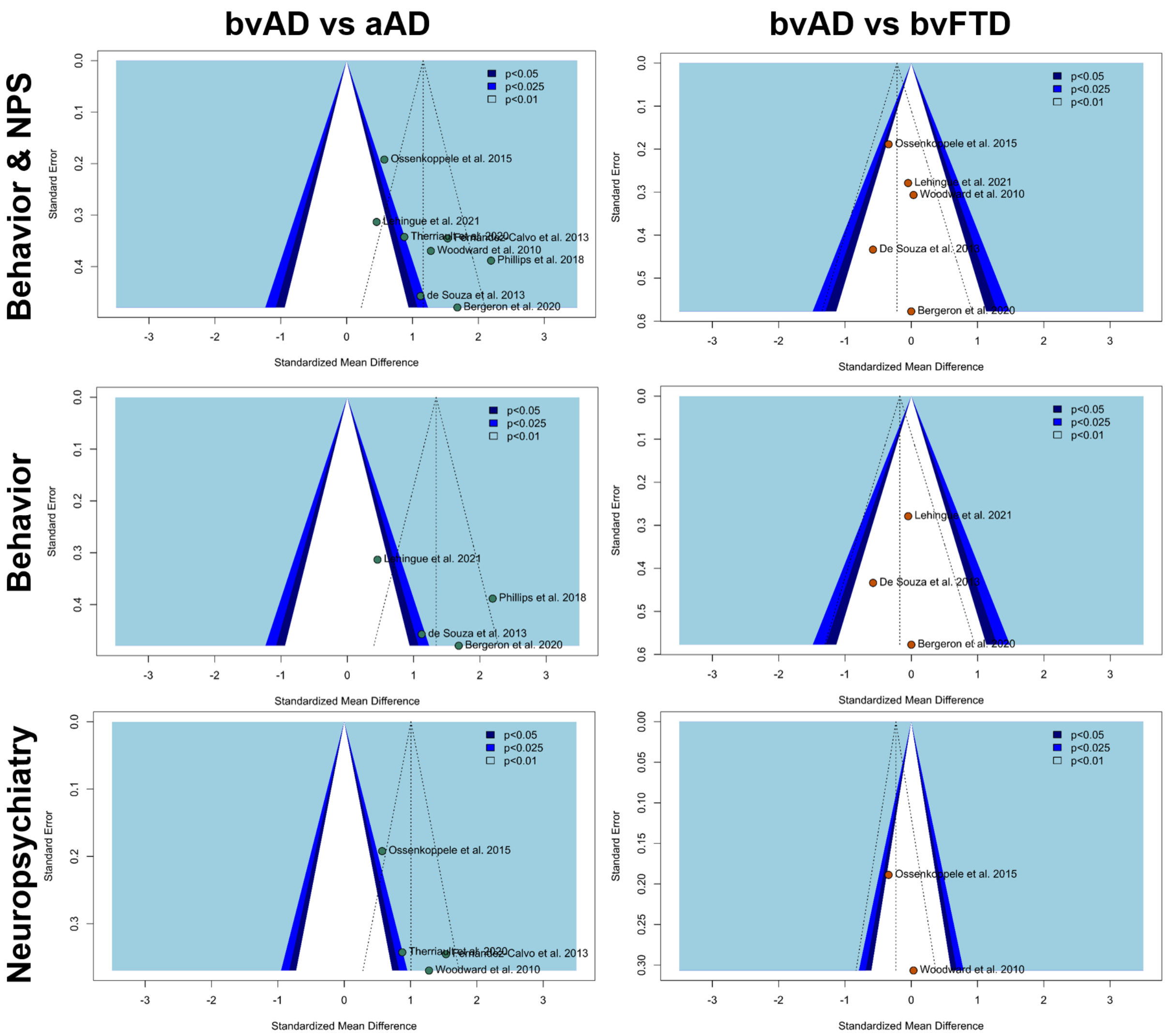


Funnel plots displaying the position of individual studies on their standardized mean difference (x-axis) relative to their standard error (y-axis). If no publication bias were present, studies would be aligned symmetrically within the dotted triangles, indicating symmetrical locations surrounding the mean effect size, with smaller studies at the lower ends of the plot and larger studies on the higher end of the plot.. The dark blue, medium dark blue and light blue parts represent the locations where the effect of the individual study is significant at p<0.05, p<0.025 and p<0.01 compared to the standardized mean difference at 0, whereas the dotted lines represent the mean effect size of the specific studies included. The current plots suggest a lower symmetrical tendency in bvAD vs tAD contrasts compared to bvAD vs bvFTD contrasts, indicating higher publication bias in the bvAD vs tAD contrasts, although the number of studies and sample sizes were small.

**Figure S2**. Funnel plots of meta-analyses for behavioral and neuropsychiatric symptom data separately for bvAD vs typical AD and bvFTD.

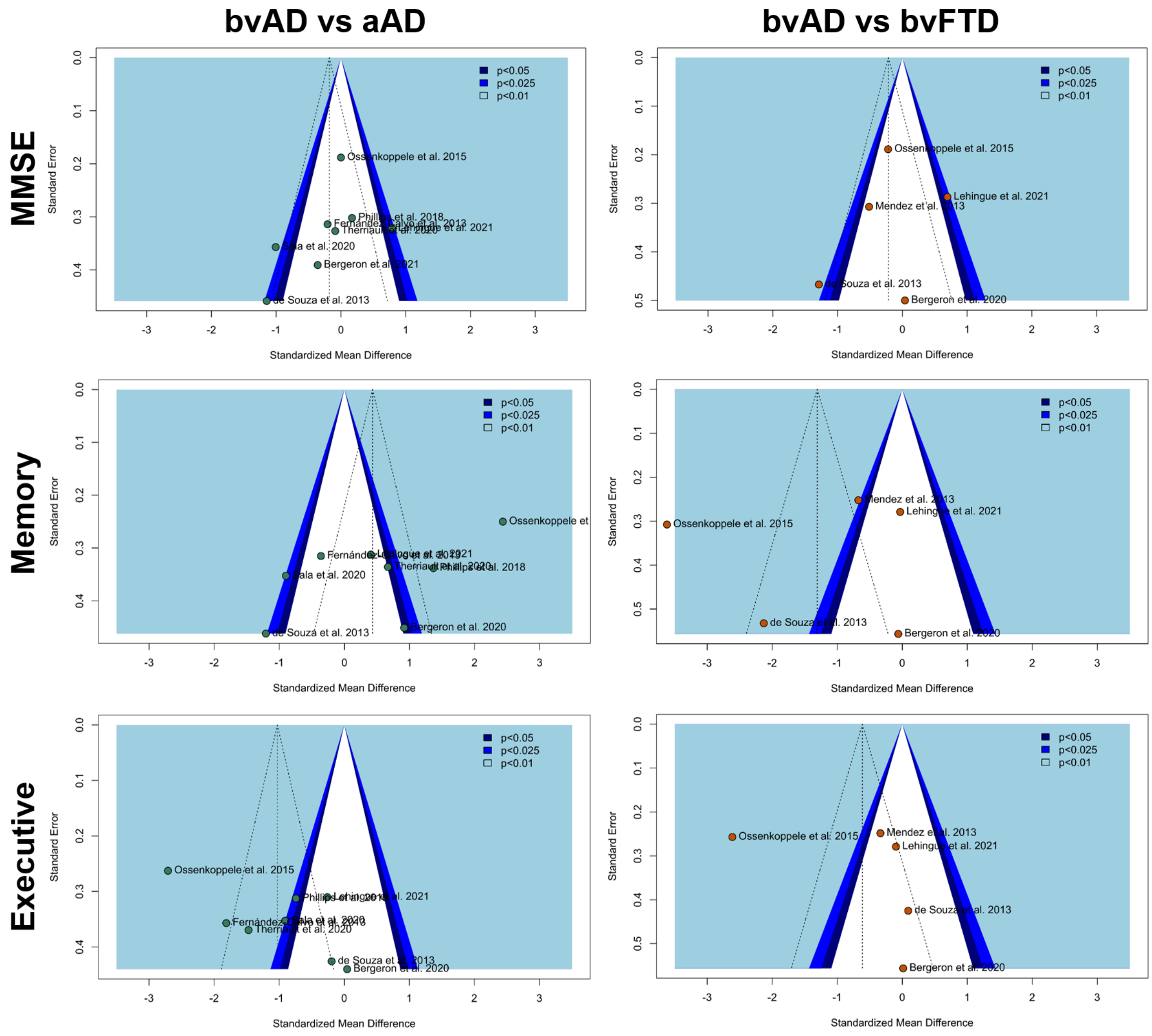


Funnel plots displaying the position of individual studies on their standardized mean difference (x-axis) relative to their standard error (y-axis). If no publication bias were present, studies would be aligned symmetrically within the dotted triangles, indicating symmetrical locations surrounding the mean effect size, with smaller studies at the lower ends of the plot and larger studies on the higher end of the plot. The dark blue, medium dark blue and light blue parts represent the locations where the effect of the individual study is significant at $p<0.05$, $p<0.025$ and $p<0.01$ compared to the standardized mean difference at 0, whereas the dotted lines represent the mean effect size of the specific studies included. The current plots suggest a higher symmetrical tendency in the MMSE contrasts than in the memory and executive domains, indicating higher publication bias in the memory and executive functioning domains than in the MMSE, although the number of studies and sample sizes were small.

**Figure S3**. Funnel plots of meta-analyses of neuropathological data in bvAD versus typical AD and bvFTD.

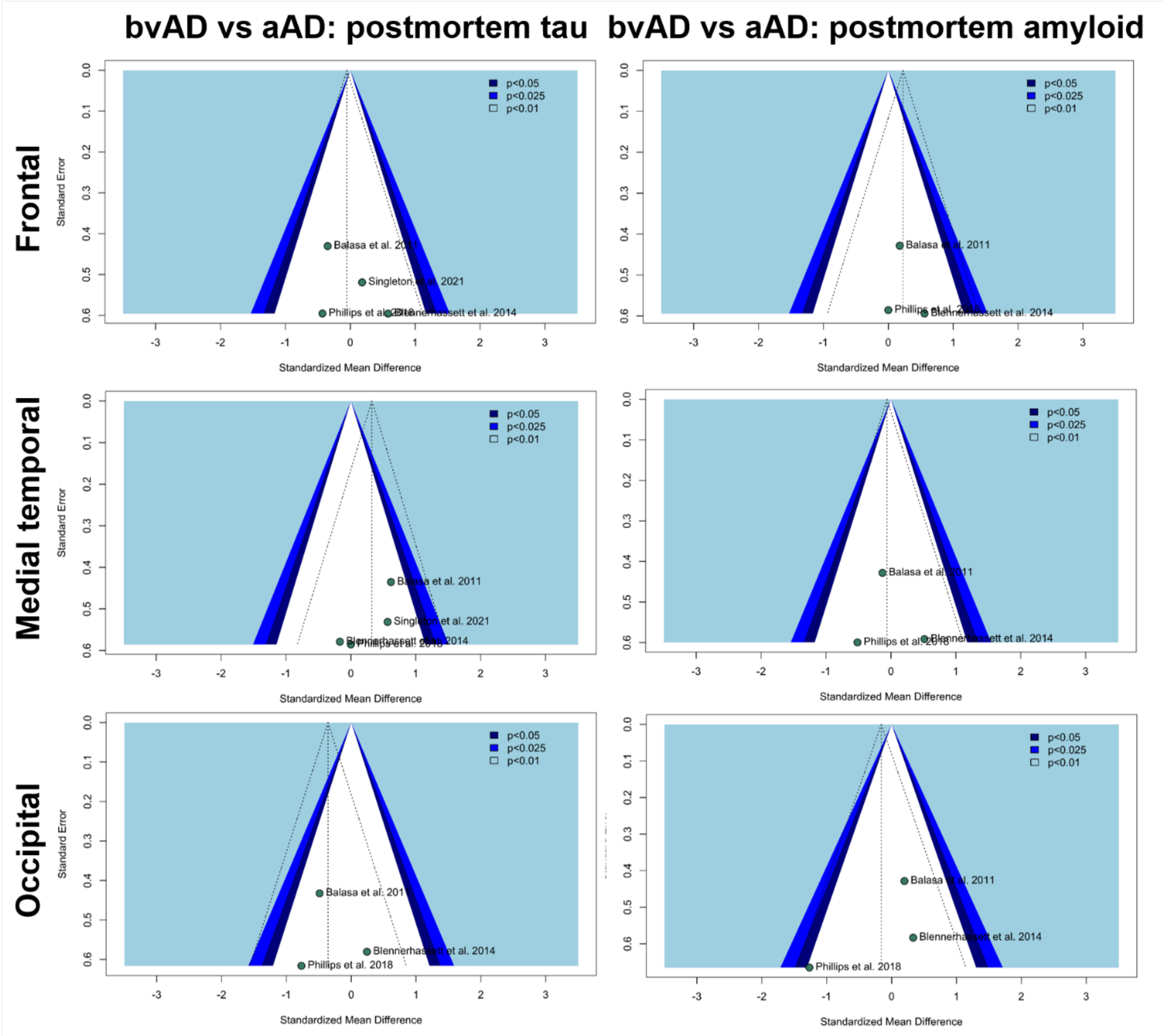


Funnel plots displaying the position of individual studies on their standardized mean difference (x-axis) relative to their standard error (y-axis). If no publication bias were present, studies would be aligned symmetrically within the dotted triangles, indicating symmetrical locations surrounding the mean effect size, with smaller studies at the lower ends of the plot and larger studies on the higher end of the plot. The dark blue, medium dark blue and light blue parts represent the locations where the effect of the individual study is significant at p<0.05, p<0.025 and p<0.01 compared to the standardized mean difference at 0, whereas the dotted lines represent the mean effect size of the specific studies included. Although few studies were included per plot, the current plots show an overall symmetrical tendency, marking marginal publication bias.

**Figure S4.** Summary results of Risk of Bias assessment according to the ROBINS-I tool for studies included in the meta-analyses

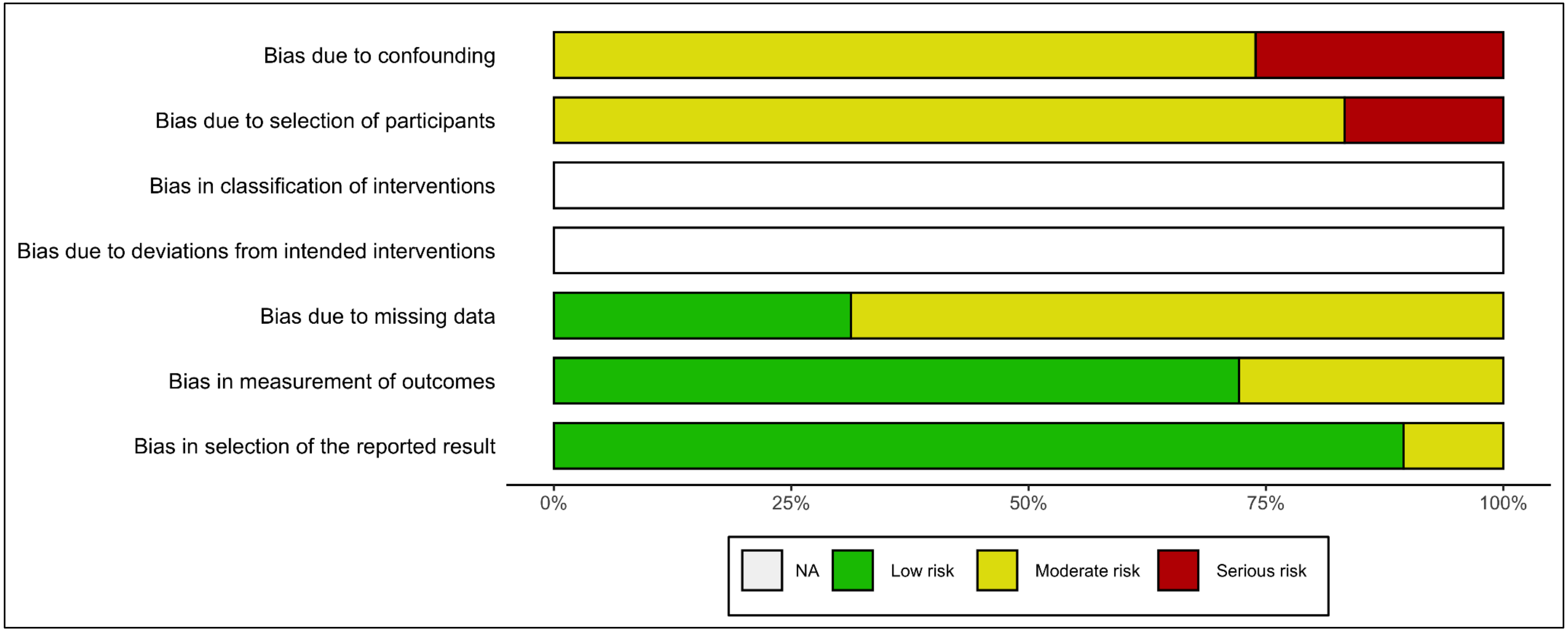


The ROBINS-I tool for non-randomized studies (https://www.riskofbias.info/) was applied to assess Risk of Bias across studies. Since the domains 'Bias in classification of interventions' and 'Bias due to deviations from intended interventions' were not applicable to the currently assessed studies, these were not filled out (*NA*=not available). See Table S3 for further details.

**Figure S5**. Flow chart of study inclusion

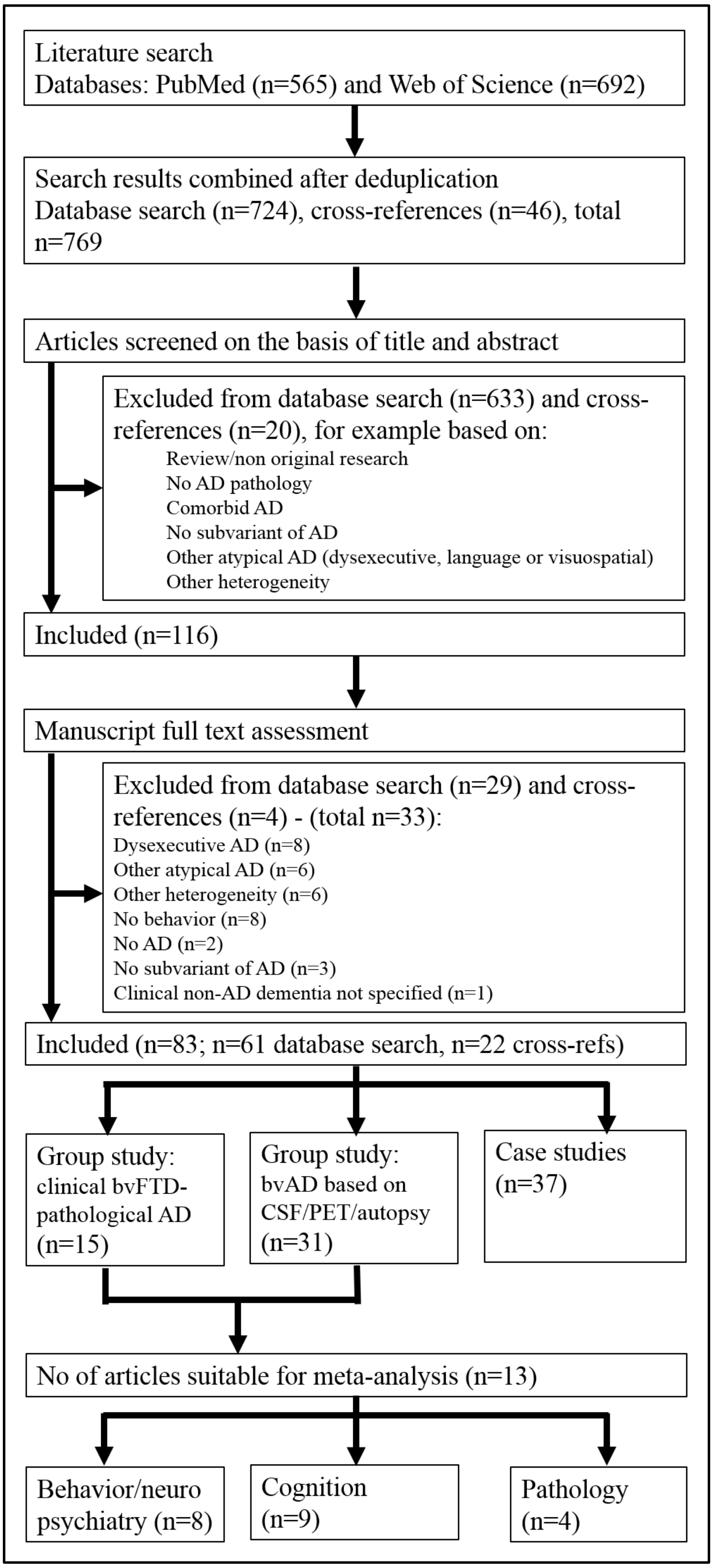


*AD*=Alzheimer's disease, *bvAD*=behavioral variant of AD, *bvFTD*=behavioral variant frontotemporal dementia, *CSF*=cerebrospinal fluid, *PET*=positron emission tomography.

**Figure S6.** Meta-analyses for behavior and neuropsychiatric symptoms separately in bvAD vs typical AD and bvFTD

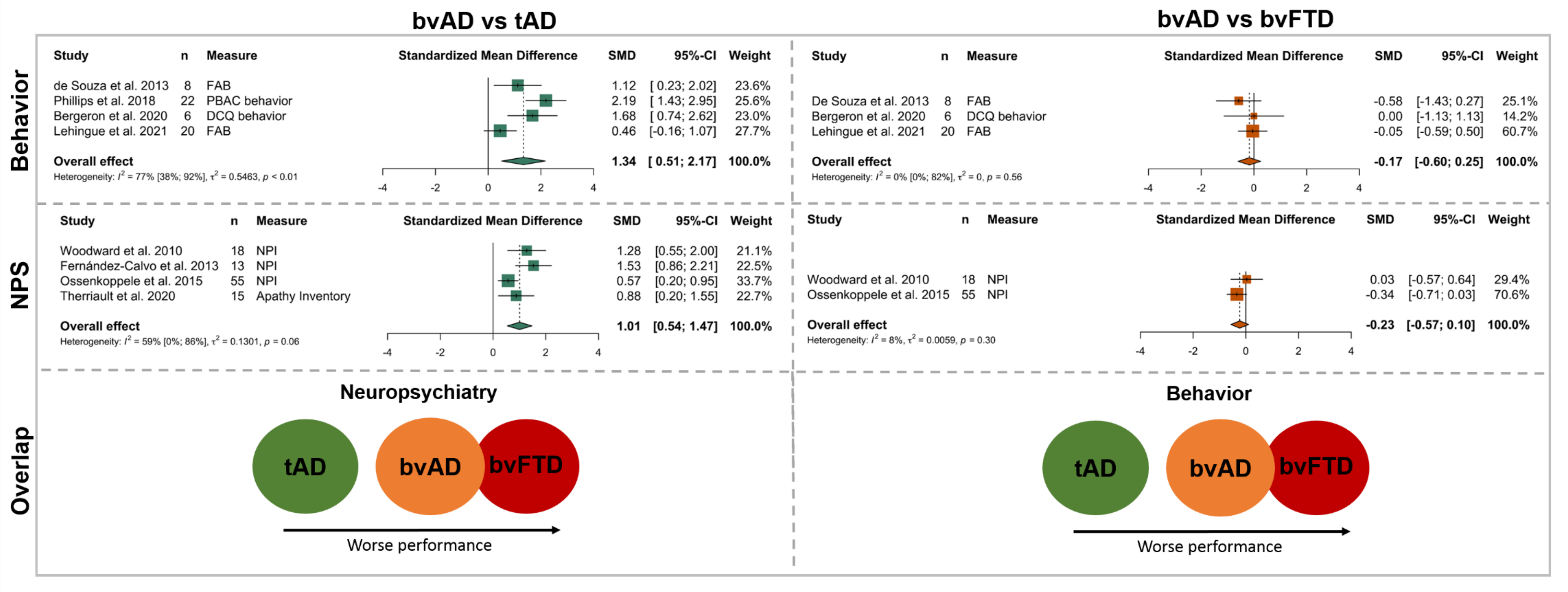


Plots showing meta-analysis results for behavior and neuropsychiatric symptoms separately between patient groups. These plots show similar scores in both behavioral as neuropsychiatric scales scores in bvAD versus bvFTD and a similar difference in behavioral and neuropsychiatric scale scores in bvAD versus typical AD. For all meta-analyses, positive standardized mean differences indicate a greater neuropathological burden in bvAD versus typical AD.

*bvAD*=behavioral variant of AD, *tAD*=typical AD, *bvFTD*=behavioral variant frontotemporal dementia, *FAB*=Frontal Assessment Battery, *PBAC*=Philadelphia Brief Assessment of Cognition, *DCQ*=Dépistage Cognitif de Québec, *SMD*=standardized mean difference.

**Figure S7.** Meta-analyses for neuropathological data in bvAD vs typical AD

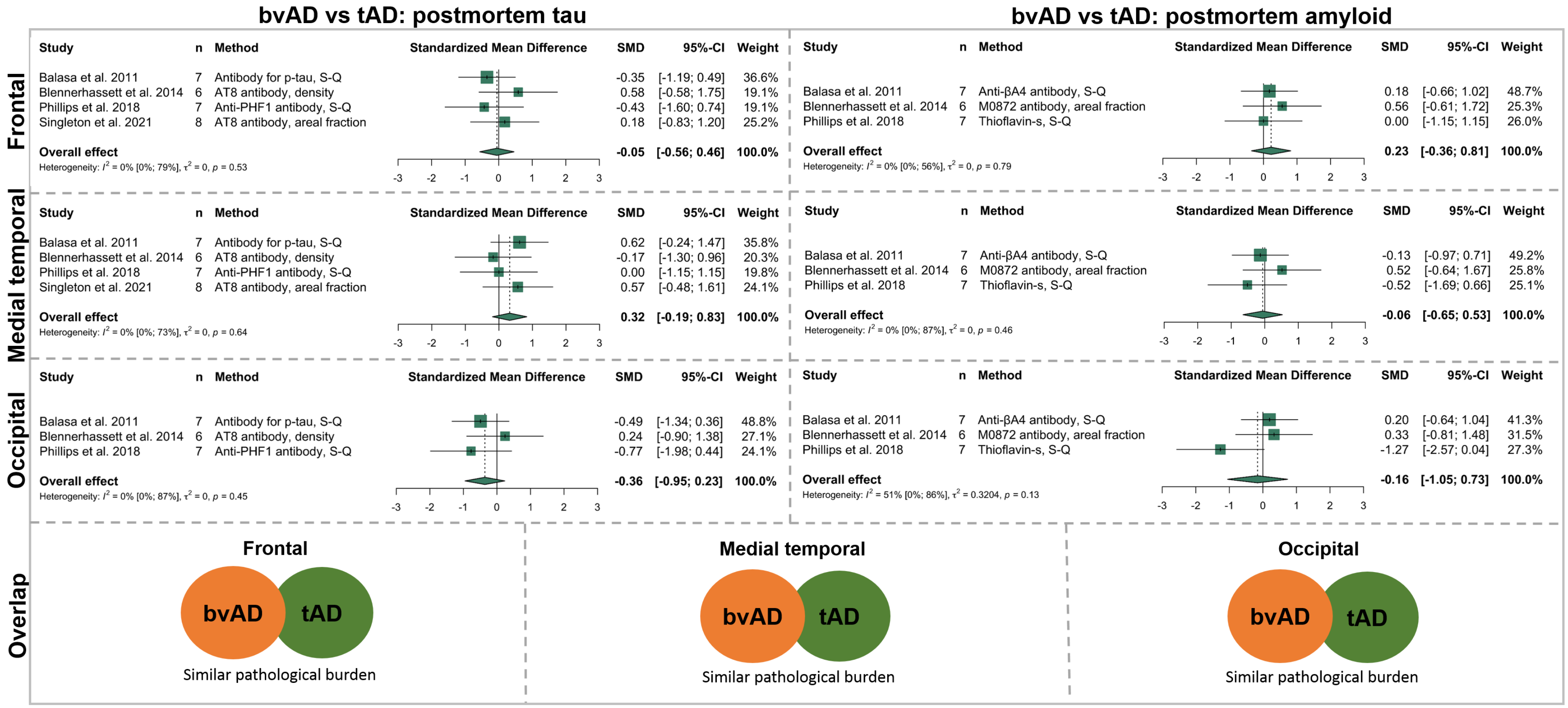


The figure shows results of meta-analyses across frontal (top row), medial temporal (middle row) and occipital (bottom row) regional quantification of postmortem tau (left column) and amyloid-β (right column) pathology in bvAD versus typical AD. For all meta-analyses, positive standardized mean differences indicate a greater neuropathological burden in bvAD versus typical AD.

*SMD*=standardized mean difference, *S-Q*=semi-quantitative.

#### REFERENCES:


1. Woodward M, Jacova C, Black SE, et al. Differentiating the frontal variant of Alzheimer's disease. *International Journal of Geriatric Psychiatry.* 2010;25(7):732-738.
2. Balasa M, Gelpi E, Antonell A, et al. Clinical features and APOE genotype of pathologically proven early-onset Alzheimer disease. *Neurology.* 2011;76(20):1720-1725.
3. de Souza LC, Bertoux M, Funkiewiez A, et al. Frontal presentation of Alzheimer's disease: a series of patients with biological evidence by CSF biomarkers. *Dementia & neuropsychologia.* 2013;7(1):66-74.
4. Mendez MF, Joshi A, Tassniyom K, Teng E, Shapira JS. Clinicopathologic differences among patients with behavioral variant frontotemporal dementia. *Neurology.* 2013;80(6):561-568.
5. Fernandez-Calvo B, Ramos F, de Lucena VM. Frontal variant of Alzheimer's disease and typical Alzheimer's disease: A comparative study. *Anales De Psicologia.* 2013;29(1):293-300.
6. Blennerhassett R, Lillo P, Halliday GM, Hodges JR, Kril JJ. Distribution of pathology in frontal variant Alzheimer's disease. *Journal of Alzheimer's disease : JAD.* 2014;39(1):63-70.
7. Ossenkoppele R, Pijnenburg YA, Perry DC, et al. The behavioural/dysexecutive variant of Alzheimer's disease: clinical, neuroimaging and pathological features. *Brain : a journal of neurology.* 2015;138(Pt 9):2732-2749.
8. Phillips JS, Da Re F, Dratch L, et al. Neocortical origin and progression of gray matter atrophy in nonamnestic Alzheimer's disease. *Neurobiol Aging.* 2018;63:75-87.
9. Sala A, Caprioglio C, Santangelo R, et al. Brain metabolic signatures across the Alzheimer's disease spectrum. *Eur J Nucl Med Mol Imaging.* 2020;47(2):256-269.
10. Therriault J, Pascoal TA, Savard M, et al. Topographical distribution of amyloid-β, tau and atrophy in behavioral / dysexecutive AD patients. *Neurology.* 2020:10.1212/WNL.0000000000011081.
11. Bergeron D, Beauregard JM, Jean G, et al. Posterior Cingulate Cortex Hypometabolism in Non-Amnestic Variants of Alzheimer's Disease. *Journal of Alzheimers Disease.* 2020;77(4):1569-1577.
12. Singleton E, Hansson O, Pijnenburg YAL, et al. Heterogeneous distribution of tau pathology in the behavioural variant of Alzheimer's disease. *Journal of Neurology, Neurosurgery & Psychiatry.* 2021:jnnp-2020-325497.
13. Lehingue E, Gueniat J, Jourdaa S, et al. Improving the Diagnosis of the Frontal Variant of Alzheimer's Disease with the DAPHNE Scale. *Journal of Alzheimers Disease.* 2021;79(4):1735-1745.
14. Brun A, Gustafson L. Distribution of cerebral degeneration in Alzheimer's disease. A clinico-pathological study. *Archiv fur Psychiatrie und Nervenkrankheiten.* 1976;223(1):15-33.
15. Shibayama H, Kitoh J. Electron microscopic structure of the Alzheimer's neurofibrillary changes in case of atypical senile dementia. *Acta Neuropathol.* 1978;41(3):229-234.
16. Shuttleworth EC. Atypical presentations of dementia of the Alzheimer type. *Journal of the American Geriatrics Society.* 1984;32(7):485-490.
17. Brun A. Frontal lobe degeneration of non-Alzheimer type. I. Neuropathology. *Archives of gerontology and geriatrics.* 1987;6(3):193-208.
18. Perani D, Di Piero V, Vallar G, et al. Technetium-99m HM-PAO-SPECT study of regional cerebral perfusion in early Alzheimer's disease. *J Nucl Med.* 1988;29(9):1507-1514.
19. Bird TD, Sumi SM, Nemens EJ, et al. Phenotypic heterogeneity in familial alzheimer's disease: A study of 24 kindreds. *Annals of Neurology.* 1989;25(1):12-25.
20. Grady CL, Haxby JV, Schapiro MB, et al. Subgroups in dementia of the Alzheimer type identified using positron emission tomography. *The Journal of neuropsychiatry and clinical neurosciences.* 1990;2(4):373-384.
21. Molchan SE, Martinez RA, Lawlor BA, Grafman JH, Sunderland T. Reflections of the self: atypical misidentification and delusional syndromes in two patients with Alzheimer's disease. *The British journal of psychiatry : the journal of mental science.* 1990;157:605-608.

22. Raux G, Gantier R, Thomas-Anterion C, et al. Dementia with prominent frontotemporal features associated with L113P presenilin 1 mutation. *Neurology.* 2000;55(10):1577-1578.
23. Rippon GA, Crook R, Baker M, et al. Presenilin 1 mutation in an african american family presenting with atypical Alzheimer dementia. *Archives of neurology.* 2003;60(6):884-888.
24. Yokota O, Terada S, Ishizu H, et al. Variability and heterogeneity in Alzheimer's disease with cotton wool plaques: a clinicopathological study of four autopsy cases. *Acta Neuropathol.* 2003;106(4):348-356.
25. Doran M, Larner AJ. Prominent behavioural and psychiatric symptoms in early-onset Alzheimer's disease in a sib pair with the presenilin-1 gene R269G mutation. *European archives of psychiatry and clinical neuroscience.* 2004;254(3):187-189.
26. Kertesz A, McMonagle P, Blair M, Davidson W, Munoz DG. The evolution and pathology of frontotemporal dementia. *Brain.* 2005;128(Pt 9):1996-2005.
27. Shi J, Shaw CL, Du Plessis D, et al. Histopathological changes underlying frontotemporal lobar degeneration with clinicopathological correlation. *Acta Neuropathol.* 2005;110(5):501-512.
28. Forman MS, Farmer J, Johnson JK, et al. Frontotemporal dementia: Clinicopathological correlations. *Annals of Neurology.* 2006;59(6):952-962.
29. Larner AJ. "Frontal variant Alzheimer's disease": a reappraisal. *Clinical neurology and neurosurgery.* 2006;108(7):705-708.
30. Alladi S, Xuereb J, Bak T, et al. Focal cortical presentations of Alzheimer's disease. *Brain.* 2007;130(Pt 10):2636-2645.
31. Rabinovici GD, Furst AJ, Neil JP, et al. 11C-PIB PET imaging in Alzheimer disease and frontotemporal lobar degeneration. *Neurology.* 2007;68(15):1205.
32. Snowden JS, Stopford CL, Julien CL, et al. Cognitive phenotypes in Alzheimer's disease and genetic risk. *Cortex; a journal devoted to the study of the nervous system and behavior.* 2007;43(7):835-845.
33. Taylor KI, Probst A, Miserez AR, Monsch AU, Tolnay M. Clinical course of neuropathologically confirmed frontal-variant Alzheimer's disease. *Nature clinical practice Neurology.* 2008;4(4):226-232.
34. Kile SJ, Ellis WG, Olichney JM, Farias S, DeCarli C. Alzheimer abnormalities of the amygdala with Klüver-Bucy syndrome symptoms: an amygdaloid variant of Alzheimer disease. *Archives of neurology.* 2009;66(1):125-129.
35. Bigio EH, Mishra M, Hatanpaa KJ, et al. TDP-43 pathology in primary progressive aphasia and frontotemporal dementia with pathologic Alzheimer disease. *Acta Neuropathol.* 2010;120(1):43-54.
36. Habek M, Hajnsek S, Zarkovic K, Chudy D, Mubrin Z. Frontal Variant of Alzheimer's Disease: Clinico-CSF-Pathological Correlation. *Canadian Journal of Neurological Sciences.* 2010;37(1):118-120.
37. Lehmann M, Rohrer JD, Clarkson MJ, et al. Reduced cortical thickness in the posterior cingulate gyrus is characteristic of both typical and atypical Alzheimer's disease. *Journal of Alzheimer's disease : JAD.* 2010;20(2):587-598.
38. Piscopo P, Talarico G, Crestini A, et al. A novel mutation in the predicted TMIII domain of the PSEN2 gene in an Italian pedigree with atypical Alzheimer's disease. *Journal of Alzheimer's disease : JAD.* 2010;20(1):43-47.
39. Rabinovici GD, Rosen HJ, Alkalay A, et al. Amyloid vs FDG-PET in the differential diagnosis of AD and FTLD. *Neurology.* 2011;77(23):2034-2042.
40. Snowden JS, Thompson JC, Stopford CL, et al. The clinical diagnosis of early-onset dementias: diagnostic accuracy and clinicopathological relationships. *Brain.* 2011;134(Pt 9):2478-2492.
41. Whitwell JL, Jack CR, Przybelski SA, et al. Temporoparietal atrophy: A marker of AD pathology independent of clinical diagnosis. *Neurobiology of Aging.* 2011;32(9):1531-1541.
42. Borroni B, Pilotto A, Bonvicini C, et al. Atypical presentation of a novel Presenilin 1 R377W mutation: sporadic, late-onset Alzheimer disease with epilepsy and frontotemporal atrophy.

*Neurological sciences : official journal of the Italian Neurological Society and of the Italian Society of Clinical Neurophysiology.* 2012;33(2):375-378.

43. Duker AP, Espay AJ, Wszolek ZK, Rademakers R, Dickson DW, Kelley BJ. Atypical motor and behavioral presentations of Alzheimer disease: a case-based approach. *The neurologist.* 2012;18(5):266-272.
44. Wallon D, Rousseau S, Rovelet-Lecrux A, et al. The French series of autosomal dominant early onset Alzheimer's disease cases: mutation spectrum and cerebrospinal fluid biomarkers. *Journal of Alzheimer's disease : JAD.* 2012;30(4):847-856.
45. Herrero-San Martin A, Villarejo-Galende A, Rabano-Gutierrez A, Guerrero-Marquez C, Porta-Etessam J, Bermejo-Pareja F. Frontal variant of Alzheimer's disease. Two pathologically confirmed cases and a literature review. *Revista De Neurologia.* 2013;57(12):542-548.
46. Marini S, Lucidi G, Tedde A, Bessi V, Nacmias B. A case of atypical early-onset Alzheimer's disease carrying the missense mutation Thr354Ile in exon 10 of the PSEN1 gene. *Neurological sciences : official journal of the Italian Neurological Society and of the Italian Society of Clinical Neurophysiology.* 2013;34(9):1691-1692.
47. Léger GC, Banks SJ. Neuropsychiatric symptom profile differs based on pathology in patients with clinically diagnosed behavioral variant frontotemporal dementia. *Dementia and geriatric cognitive disorders.* 2014;37(1-2):104-112.
48. Nygaard HB, Lippa CF, Mehdi D, Baehring JM. A Novel Presenilin 1 Mutation in Early-Onset Alzheimer's Disease With Prominent Frontal Features. *American journal of Alzheimer's disease and other dementias.* 2014;29(5):433-435.
49. Balasa M, Gelpi E, Martín I, et al. Diagnostic accuracy of behavioral variant frontotemporal dementia consortium criteria (FTDC) in a clinicopathological cohort. *Neuropathology and applied neurobiology.* 2015;41(7):882-892.
50. Paterson RW, Toombs J, Slattery CF, et al. Dissecting IWG-2 typical and atypical Alzheimer's disease: insights from cerebrospinal fluid analysis. *J Neurol.* 2015;262(12):2722-2730.
51. Woodward MC, Rowe CC, Jones G, Villemagne VL, Varos TA. Differentiating the frontal presentation of Alzheimer's disease with FDG-PET. *Journal of Alzheimer's disease : JAD.* 2015;44(1):233-242.
52. Li P, Zhou YY, Lu D, Wang Y, Zhang HH. Correlated patterns of neuropsychological and behavioral symptoms in frontal variant of Alzheimer disease and behavioral variant frontotemporal dementia: a comparative case study. *Neurological sciences : official journal of the Italian Neurological Society and of the Italian Society of Clinical Neurophysiology.* 2016;37(5):797-803.
53. Ossenkoppele R, Schonhaut DR, Scholl M, et al. Tau PET patterns mirror clinical and neuroanatomical variability in Alzheimer's disease. *Brain : a journal of neurology.* 2016;139(Pt 5):1551-1567.
54. Scialò C, Ferrara M, Accardo J, et al. Frontal Variant Alzheimer Disease or Frontotemporal Lobe Degeneration With Incidental Amyloidosis? *Alzheimer Dis Assoc Disord.* 2016;30(2):183-185.
55. Dickerson BC, McGinnis SM, Xia C, et al. Approach to atypical Alzheimer's disease and case studies of the major subtypes. *CNS spectrums.* 2017;22(6):439-449.
56. Duclos H, de La Sayette V, Bonnet AL, et al. Social Cognition in the Frontal Variant of Alzheimer's Disease: A Case Study. *Journal of Alzheimer's disease : JAD.* 2017;55(2):459-463.
57. Kawakatsu S, Kobayashi R, Hayashi H. Typical and atypical appearance of early-onset Alzheimer's disease: A clinical, neuroimaging and neuropathological study. *Neuropathology : official journal of the Japanese Society of Neuropathology.* 2017;37(2):150-173.
58. Oboudiyat C, Gefen T, Varelas E, et al. Cerebrospinal fluid markers detect Alzheimer's disease in nonamnestic dementia. *Alzheimers Dement.* 2017;13(5):598-601.
59. Perry DC, Brown JA, Possin KL, et al. Clinicopathological correlations in behavioural variant frontotemporal dementia. *Brain.* 2017;140(12):3329-3345.

60. Rawtaer I, Krishnamoorthy A. Co-occurring frontal variant Alzheimer's dementia and carrier of Huntington's disease allele with reduced penetrance. *Psychogeriatrics : the official journal of the Japanese Psychogeriatric Society.* 2017;17(6):488-490.
61. Sawyer RP, Rodriguez-Porcel F, Hagen M, Shatz R, Espay AJ. Diagnosing the frontal variant of Alzheimer's disease: a clinician's yellow brick road. *Journal of clinical movement disorders.* 2017;4:2.
62. Bagyinszky E, Lee HM, Van Giau V, et al. PSEN1 p.Thr116Ile Variant in Two Korean Families with Young Onset Alzheimer's Disease. *International journal of molecular sciences.* 2018;19(9).
63. Boon BDC, Hoozemans JJM, Lopuhaä B, et al. Neuroinflammation is increased in the parietal cortex of atypical Alzheimer's disease. *Journal of Neuroinflammation.* 2018;15(1):170.
64. Seo SW, Thibodeau M-P, Perry DC, et al. Early vs late age at onset frontotemporal dementia and frontotemporal lobar degeneration. *Neurology.* 2018;90(12):e1047-e1056.
65. Whitwell JL, Graff-Radford J, Tosakulwong N, et al. Imaging correlations of tau, amyloid, metabolism, and atrophy in typical and atypical Alzheimer's disease. *Alzheimers Dement.* 2018;14(8):1005-1014.
66. de Souza LC, Mariano LI, de Moraes RF, Caramelli P. Behavioral variant of frontotemporal dementia or frontal variant of Alzheimer's disease? A case study. *Dementia & neuropsychologia.* 2019;13(3):356-360.
67. Foiani MS, Cicognola C, Ermann N, et al. Searching for novel cerebrospinal fluid biomarkers of tau pathology in frontotemporal dementia: an elusive quest. *J Neurol Neurosurg Psychiatry.* 2019;90(7):740-746.
68. Monacelli F, Martella L, Parodi MN, Odetti P, Fanelli F, Tabaton M. Frontal Variant of Alzheimer's Disease: A Report of a Novel PSEN1 Mutation. *Journal of Alzheimer's disease : JAD.* 2019;70(1):11-15.
69. Nolan A, Resende EDF, Petersen C, et al. Astrocytic Tau Deposition Is Frequent in Typical and Atypical Alzheimer Disease Presentations. *Journal of Neuropathology and Experimental Neurology.* 2019;78(12):1112-1123.
70. Pawlowski M, Joksch V, Wiendl H, Meuth SG, Duning T, Johnen A. Apraxia screening predicts Alzheimer pathology in frontotemporal dementia. *J Neurol Neurosurg Psychiatry.* 2019;90(5):562-569.
71. Phillips JS, Da Re F, Irwin DJ, et al. Longitudinal progression of grey matter atrophy in non-amnestic Alzheimer's disease. *Brain.* 2019;142(6):1701-1722.
72. Pillai JA, Bonner-Jackson A, Bekris LM, Safar J, Bena J, Leverenz JB. Highly Elevated Cerebrospinal Fluid Total Tau Level Reflects Higher Likelihood of Non-Amnestic Subtype of Alzheimer's Disease. *Journal of Alzheimer's disease : JAD.* 2019;70(4):1051-1058.
73. Tan RH, Yang Y, McCann H, Shepherd C, Halliday GM. Von Economo Neurons in Behavioral Variant Frontotemporal Dementia with Underlying Alzheimer's Disease. *Journal of Alzheimer's disease : JAD.* 2019;69(4):963-967.
74. Wang Y, Shi Z, Zhang N, et al. Spatial Patterns of Hypometabolism and Amyloid Deposition in Variants of Alzheimer's Disease Corresponding to Brain Networks: a Prospective Cohort Study. *Molecular imaging and biology.* 2019;21(1):140-148.
75. Wong S, Strudwick J, Devenney E, Hodges JR, Piguet O, Kumfor F. Frontal variant of Alzheimer's disease masquerading as behavioural-variant frontotemporal dementia: a case study comparison. *Neurocase.* 2019;25(1-2):48-58.
76. Cai H, Ning S, Li W, Li X, Xiao S, Sun L. Patient with frontal-variant syndrome in early-onset Alzheimer's disease. *General psychiatry.* 2020;33(2):e100173.
77. Cousins KAQ, Irwin DJ, Wolk DA, et al. ATN status in amnestic and non-amnestic Alzheimer's disease and frontotemporal lobar degeneration. *Brain.* 2020;143(7):2295-2311.
78. Li CH, Fan SP, Chen TF, Chiu MJ, Yen RF, Lin CH. Frontal variant of Alzheimer's disease with asymmetric presentation mimicking frontotemporal dementia: Case report and literature review. *Brain and behavior.* 2020;10(3):e01548.

79. Paquin V, Therriault J, Pascoal TA, Rosa-Neto P, Gauthier S. Frontal Variant of Alzheimer Disease Differentiated From Frontotemporal Dementia Using in Vivo Amyloid and Tau Imaging. *Cognitive and behavioral neurology : official journal of the Society for Behavioral and Cognitive Neurology.* 2020;33(4):288-293.
80. Scarioni M, Gami-Patel P, Timar Y, et al. Frontotemporal Dementia: Correlations Between Psychiatric Symptoms and Pathology. *Annals of Neurology.* 2020;87(6):950-961.
81. Singleton EH, Pijnenburg YAL, Sudre CH, et al. Investigating the clinico-anatomical dissociation in the behavioral variant of Alzheimer disease. *Alzheimers Res Ther.* 2020;12(1):148.
82. Bergeron D, Sellami L, Poulin S, Verret L, Bouchard RW, Laforce R. The Behavioral/Dysexecutive Variant of Alzheimer's Disease: A Case Series with Clinical, Neuropsychological, and FDG-PET Characterization. *Dementia and Geriatric Cognitive Disorders.* 2021;49(5):518-525.
83. Zhu L, Sun LM, Sun L, Xiao SF. Case of early-onset Alzheimer's disease with atypical manifestation. *General Psychiatry.* 2021;34(1).
84. Eikelboom WS, van den Berg E, Singleton EH, et al. Neuropsychiatric and Cognitive Symptoms Across the Alzheimer Disease Clinical Spectrum: Cross-sectional and Longitudinal Associations. *Neurology.* 2021.
